# SAFETY AND IMMUNOGENICITY OF A PHH-1V BOOSTER DOSE AFTER DIFFERENT PRIME VACCINATION SCHEMES AGAINST COVID-19: PHASE III CLINICAL TRIAL FINAL RESULTS UP TO ONE YEAR

**DOI:** 10.1101/2024.05.14.24307343

**Authors:** Silvina Natalini Martínez, Rafael Ramos, Jorge Navarro-Perez, Maria Jesus Lopez, Maria del Mar Vazquez, Jose Molto, Patricia Munoz, Jose M Echave, Jose R Arribas, Melchor Alvarez, Eunate Arana-Arri, Jordi Calls, Susana Otero-Romero, Francesco Scaglione, Laia Bernad, Raúl Pérez-Caballero, Julia G Prado, Ignasi Esteban, Elena Aurrecoechea, Roc Pomarol, Montserrat Plana, Alex Soriano

## Abstract

In this phase III, open label, single arm, multicenter clinical study, we report safety, tolerability and immunogenicity of PHH-1V as a booster dose in subjects primary vaccinated against COVID-19 with the BNT162b2, mRNA-1273, ChAdOx1-S, or Ad26.COV2.S vaccines, with or without previous COVID-19 infection. A total of 2661 subjects were included in this study and vaccinated with the PHH-1V vaccine. Most treatment-emergent adverse events (TEAE) were solicited local and systemic reactions with grade 1 (58.70%) or grade 2 (27.58%) intensity, being the most frequently reported injection site pain (82.83%), fatigue (31.72%) and headache (31.23%). Additionally, immunogenicity was assessed at Baseline and Days 14, 91, 182 and 365 in a subset of 235 subjects primary vaccinated. On Day 14, geometric mean triter (GMT) in neutralizing antibody against SARS-CoV-2 Wuhan and Beta, Delta and Omicron BA.1 variants increased in all primary vaccination with a geometric mean fold raise (GMFR) of 6.90 (95% CI 4.96-9.58), 12.27 (95% CI 8.52-17.67), 7.24 (95% CI 5.06-10.37) and 17.51 (95% CI 12.28-24.97), respectively. Despite GMT decay after day 14, it remains in all cases significatively higher from baseline up to 1 year after PHH-1V booster administration and GMFR against Beta and Omicron BA.1 variants over 3 at 1 year after booster compared to baseline. PHH-1V booster vaccination elicited also a significant RBD/Spike-specific IFN-γ^+^ T-cell responses on Day 14. Overall, PHH-1V vaccine was immunogenic and well-tolerated regardless of the previous primary vaccination scheme received with no reported cases of severe COVID-19 infection throughout the entire study.

## INTRODUCTION

Coronavirus disease 2019 (COVID-19), caused by severe acute respiratory syndrome coronavirus 2 (SARS-CoV-2), remains a global health issue with a total of 774 million confirmed cases and more than 7 million deaths reported globally as of March 2024^1^. Although COVID-19 vaccines marketed for human use dramatically reduced hospitalization and mortality^2,3^, global vaccine supply is inequitable and only 32.7% of people in low-income countries have received at least one dose^4^. Moreover, several studies have reported a decline in the effectiveness of COVID-19 vaccines in preventing symptomatic and severe disease over time^5–8^. For instance, a meta-analysis estimated that effectiveness was reduced around 10% for severe disease and around 20% for infection over 5 months^9^. This waning of protection has also been enhanced by the emergence of new variants, characterized by increased immune evasion and transmissibility in vaccinated individuals^10,11^. This situation has underlined the need of novel vaccines for booster immunization to supplement the initial primary vaccination and maintain adequate protection of the population. It has been reported that booster doses enhance immunogenicity and restore protection against the Omicron variant^12–15^. The European Centre for Disease Prevention and Control (ECDC) and the European Medicines Agency (EMA) on winter season of 2023 recommended additional booster doses with adapted COVID-19 vaccines in people aged 60 years and above, the immunocompromised, and other vulnerable persons (from 12 years of age) with underlying conditions putting them at higher risk of severe COVID-19, and pregnant women^16^.

Several adapted bivalent and monovalent COVID-19 booster vaccines have been developed to fulfil the demanding vaccine situation and to immunize primary vaccinated individuals against the emerging variants. PHH-1V (BIMERVAX^®^; HIPRA S.A.) is a recombinant bivalent heterodimer adjuvanted vaccine for the prevention of COVID-19, prepared as an emulsion for intramuscular administration in individuals 16 years old and older. The vaccine antigen consists of a fusion heterodimer of 2 receptor binding domain (RBD) from the SARS-CoV-2 Beta (B.1.351) and Alpha (B.1.1.7) variants produced as a single chain recombinant antigen in Chinese Hamster Ovary (CHO) cells and adjuvanted with an oil-in-water emulsion based on squalene (SQBA). Subunit adjuvanted vaccines based on recombinant proteins offer several advantages since they can be efficiently produced on expression platforms and scaled easily at high yields, making them easier to produce and distribute globally. They are also stable and less reliant on a cold chain for their distribution than other COVID-19 vaccines^19^.

A first-in-human Phase I/IIa dose-escalation, randomized, double-blinded, active-comparator controlled clinical trial in 30 healthy adults demonstrated that two doses of PHH-1V vaccine in a range of 10 to 40 µg/dose were safe and well-tolerated and induced robust humoral immune responses to different circulating variants of concern at the time of the study, including Alpha (B1.1.7), Beta (B.1.351), Delta (B.1.617.2) and Gamma (P.1)^20^ A multicenter, randomized, active-controlled, double-blind, non-inferiority Phase IIb clinical trial also showed that PHH-1V displayed a good safety profile, with fewer reported solicited adverse events compared to Pfizer/BioNTech BNT162b2 vaccine^21^. In this phase IIb study, PHH-1V vaccine elicited a powerful neutralizing antibody response against SARS-CoV-2 Beta, Delta and Omicron strains, including Omicron XBB.1.5^22^ and these results were statistically superior compared to BNT162b2 vaccine at several time points for Beta, Delta and Omicron BA.1 and non-inferiority against Omicron XBB.1.5 ^21,22^. Moreover, the PHH-1V boost also induced a strong and sustained T-cell response against different SARS-CoV-2 variants^21,22^.

Here, we report the results of a Phase III, open label, single arm, multicenter clinical study in healthy adults primary vaccinated against COVID-19 (NCT05303402). This Phase III clinical study aimed to assess the safety and reactogenicity of a booster dose of PHH-1V vaccine as a heterologous booster in subjects who had been vaccinated (following local authorities recognized vaccination scheme) at least 91 days before administration of study vaccine. Participants who had previously suffered from a non-severe or asymptomatic COVID-19 infection were also included. Immunogenicity was also assessed in a subset of subjects previously vaccinated either with homologous or heterologous schemes with BNT162b2 (Comirnaty; Pfizer/BioNTech), ChAdOx1-S (Vaxzevria; AstraZeneca) or mRNA-1273 (Spikevax; Moderna) vaccines.

## MATERIALS AND METHODS

### Study design and participants

This was a Phase III, open label, single arm, multicenter clinical trial to assess safety, reactogenicity, tolerability and immunogenicity of a booster vaccination with PHH-1V, a recombinant protein RBD fusion heterodimer vaccine against COVID-19. The study was conducted at 17 study centers in Spain and 1 center in Italy.

Eligible participants were individuals aged 16 years or older who had a COVID-19 vaccination scheme recognized by the authorities of the country with BNT162b2 (Comirnaty; Pfizer/BioNTech), ChAdOx1-S (Vaxzevria; AstraZeneca), mRNA-1273 (Spikevax; Moderna) or Ad26.COV2-S (Jcovden; Janssen) at least 91 days (preferably a maximum of 240 days) before Day 0; provided written informed consent, and were agreed not to donate blood, blood products and bone marrow at least 3 months before and after vaccination. For the safety assessment, subjects completed a primary vaccination schedule with the vaccines mentioned above (including those suffering non-severe COVID-19 after second dose at least 30 days before Day 0) or were vaccinated with one dose and suffered non-severe COVID-19 infection (confirmed by RT-qPCR or rapid antigen test) before or after receiving the one dose. For immunogenicity assessment, subjects were primary vaccinated with one of the following vaccination schemes: two doses of ChAdOx1-S, two doses of mRNA-1273 or one dose of ChAdOx1-S combined with an mRNA vaccine and without previous SARS-CoV-2 infection. In subjects aged 16 or 17 years old, only those primary vaccinated with two doses of BNT162b2 with no previous COVID-19 infection were eligible for immunogenicity assessment. Additionally, female subjects of childbearing potential had a negative pregnancy test on the day of vaccination and agreed to use any acceptable contraceptive method from Day 0 to 8 weeks after vaccination. Key exclusion criteria included history of anaphylaxis to any prior vaccine, previous severe SARS-CoV-2 infection, previous immunization with live attenuated vaccines within 4 weeks before or after receiving any study vaccine, pregnant or breast-feeding at screening, clinically significant acute illness at screening or within 48 hours prior to study vaccination, surgery requiring hospitalization before vaccination, severe and non-stable psychiatric condition, and abnormal function of the immune system.

This study was conducted in accordance with the ethical principles stated in the Declaration of Helsinki and in the International Council for Harmonization (ICH) guidelines for Good Clinical Practice (GCP), and all applicable local laws and regulations. The study protocol, informed consent form (ICF), and written information given to subjects were reviewed and approved by an appropriately constituted Institutional Review Board (IRB) and the Independent Ethics Committee (IEC) from Spain and Italy (IEC Hospital Clínic de Barcelona, and IEC LAZZARO SPALLANZANI, IRCCS). The protocol was also reviewed and approved by Spanish Agency of Medicines and Medical Devices (AEMPS), and also by Agenzia Italiana del Farmaco (AIFA). A Data Safety Monitoring Board (DSMB) was also available to review safety data at any time point during the study. Written informed consent was obtained from all participants before enrolment. The method of obtaining and documenting informed consent and the contents of the consent complied with ICH-GCP and all applicable regulatory requirements. All subject identities were kept confidential though the assignment of a unique subject number.

### Trial Procedures

This study consists of a maximum of 28-day Pre-Screening Period (by phone) prior to the Screening/Vaccination Visit, and a Follow up Period of 182 days in the safety assessment subset and 365 days in the immunogenicity assessment subset. The study visits were scheduled on Day 0, Day 14, Day 91, Day 182 and Day 365 or early termination visit (ETV). PHH-1V was supplied in a vial containing 10 doses of 0.5 mL (40 µg) ready to use and stored in a refrigerator at 2-8°C. All eligible subjects received a PHH-1V booster dose on Day 0, administered in a volume of 0.5 mL (40 µg) by intramuscular injection into the deltoid muscle. Participants were also given a hard copy diary on Day 0 to record local and systemic solicited reactions within the 7 days after vaccination. Blood samples were obtained for safety and immunogenicity assessments according to protocol schedule (**Table S1**).

### Safety assessment

Physical examinations and vital signs were recorded at each study visit before collecting blood samples, and included pulse rate, blood pressure, body temperature, and oxygen saturation. Participants were observed for at least 30 min after each injection to identify any immediate adverse event (AE). Solicited local and systemic reactions were recorded daily by the participants in a subject diary until 7 days post-vaccination. Solicited local reactions included pain, tenderness, erythema/ redness and induration/ swelling. Solicited systemic reactions included fever, chills, nausea/ vomiting, malaise/ muscle pain, diarrhoea, headache, fatigue and joint pain. All subjects were provided with a Subject Diary to register all the local and systemic reactions from the time of vaccination until 7 days post-vaccination; the diary was collected at the Day 14 visit. Treatment-emergent adverse events (TEAE) including the unsolicited local and systemic AEs were reported through Day 28 after vaccination. Additionally, serious adverse events (SAEs), including suspected unexpected serious adverse reactions (SUSAR), were collected through the end of the study. AEs were coded by preferred term (PT) and system organ class (SOC) using the Medical Dictionary for Regulatory Activities (MedDRA version 26.0). An assessment of intensity was made using the general categorical descriptors outlined in the toxicity grading scale for healthy adult and adolescent subjects enrolled in preventive vaccine clinical studies ^23^. Grade 3 and 4 changes from Baseline in safety laboratory parameters on Day 14 after vaccination as well as SARS-CoV-2 infections and severe cases of COVID-19 were also reported. SARS-CoV-2 infections were assessed with a rapid antigen test or RT-qPCR following the standard procedures in the health system. Severe cases were considered as any episode of COVID-19 requiring ≥ 24 hours of hospitalization. Severe COVID-19 cases which met seriousness criteria were reported as SAEs. Non-severe COVID-19 cases were considered AEs but only confirmed COVID-19 cases occurring after ≥ 14 days post-booster were considered in the exploratory endpoints’ analysis.

### Humoral immunity assays

Neutralizing antibody titers against SARS-CoV-2 Wuhan (original sequence),Beta, Delta and Omicron BA.1 variants were determined by a pseudoviruses-based neutralization assay (PBNA) at HIPRA (Girona, Spain) using an HIV reporter pseudovirus that expresses the S protein of SARS-CoV-2 and luciferase as described previously ^24^. Neutralization capacity of the serum samples was calculated by comparing the experimental relative luminescence units (RLU) calculated from infected cells treated with each serum to the maximal RLUs (maximal infectivity calculated from untreated infected cells) and minimal RLUs (minimal infectivity calculated from uninfected cells) and expressed as percent neutralization: Neutralization (%) = (RLUmax–RLUexperimental)/(RLUmax–RLUmin) * 100. Inhibitory concentration 50 (IC_50_) values were calculated by plotting and fitting neutralization values and the log of serum dilution to a 4-parameters equation in Prism 9.0.2 (GraphPad Software, USA); and were expressed as reciprocal concentration for each individual sample and geometric mean titer (GMT) for descriptive statistics analysis at Baseline and on Day 14, Day 91, Day 182 and Day 365. Geometrical mean fold rise (GMFR) in neutralizing antibodies titer from baseline to Day 14, Day 91, Day 182 or Day 365 were also calculated.

Antibody binding immunity against the SARS-CoV-2 receptor binding domain (RBD) was assessed by Elecsys Anti-SARS-CoV-2 S immunoassay (Roche Diagnostics)according to manufacturer’s instructions. GMT at baseline, Day 14, Day 91, Day 182 and Day 365; and GMFR from baseline to Day 14, Day 91, Day 182 or Day 365 were also calculated for descriptive statistics.

### Cellular immunity assays

T-cell mediated immune responses against SARS-CoV-2 was assessed at baseline and Day 14 after vaccination by measurement of peripheral blood mononuclear cells (PBMCs) stimulation by IFN-γ enzyme-linked immune absorbent spot (IFN-γ ELISpot) and intracellular cytokine staining (ICS) in a subset of 24 subjects with two doses of ChAdOx1-S vaccine and no infection of COVID-19.

For ELISpot cryopreserved PBMCs were thawed in in FBS, then washed wit RPMI followed with a 4h incubation with RPMI complemented with 20% FBS. . Cells were counted and plated in a 96-wells ELISpot plate, previously coated with an anti-IFN-γ capture antibody, using a total of 0.2×10^6^ cells per well. Next, PBMCs were stimulated with six peptide pools of overlapping SARS-CoV-2 peptides, each encompassing the SARS-CoV-2 region S (2 pools) and RBD (4 pools covering SARS-CoV-2 Wuhan-Hu-1 strain, and alpha, beta and delta variants), described in **Table S2,** at a final concentration of 2.5 μg/mL per individual peptide pool. PBMCs were incubated at a final concentration of 2.5 μg/mL per individual peptide pool. PHA (Sigma-Aldrich; San Luis, Misouri, USA) was also used as positive control. After overnight incubation, cells were washed 6 times with PBS, incubated with biotin plus anti-human IFN-γ during 1 hour at room temperature, and washed 6 times with PBS followed by another 1-hour incubation at room temperature with streptavidin. Wells were then incubated with developing solution, followed by 10 minutes at room temperature with 0.05% Tween 20 in PBS 1X and 6 washes with tap water. Spots were counted in a CTL reader system. The total ELISpot responses were reported as the mean value of spot-forming cells per 10^6^ PBMC (SFC/10^6^ PBMC) upon stimulation with each peptide pool, after subtraction of background.

Likewise, the frequency of CD4^+^ and CD8^+^ T-cells expressing IFN-γ, interleukin-2 (IL-2), and interleukin-4 (IL-4) was assessed by ICS. Cryopreserved PBMCs were thawed in RPMI complemented medium 20% FBS and then washed two times with RPMI 10% FBS. Cells were counted and plated in a 96-wells round bottom plate using a total of 0.5×10^6^ cells per well. PBMCs were also incubated with the different peptide pools described above in the presence of 2 μg/mL of monoclonal antibodies against human CD28 (clone L293, BD Pharmingen, catalog number 340450) and CD49d (clone L25, BD Pharmingen, catalog number 340976) for 6 hours. During the last 4 hours of incubation, GolgiPlug (Brefeldin A, BD Cytofix/Cytoperm Plus, BD Bioscience, catalog number 555028) was added to block cytokine transport. After incubation, PBMCs were washed with PBS 1X + 0.5% BSA + 0.1% sodium azide and incubated for 20 minutes with FcR Blocking Reagent (Milteny Biotec, catalog number 130-059-901), then washed and stained for 25 minutes with the Live/Dead probe (LIVE/DEAD fixable near IR, Thermo Fisher Scientific, catalog number <u>L34975</u>) to discriminate dead cells as well as with surface antigens using the following antibodies: CD3-PerCP (SIK7, BD Biosciences, catalog number 345766), CD4-BV421 (clone RPA-T4, BD Horizon, catalog number 562424), CD8-BV510 (clone SK1, BD Horizon, catalog number 563919). Afterward, cells were washed twice in PBS 1X + 0.5% BSA + 0.1% sodium azide, fixed and permeabilized with Fix/Perm kit (BD) for intracellular cytokine staining. Cells were incubated then for 25 minutes with FcR Blocking Reagent (Milteny Biotec, catalog number 130-059-901), washed and stained with anti-human antibodies of IFN-γ-APC (clone 27, BD Pharmingen, catalog number 554707), IL-2-PE (clone 5344.111, BD FastImmune, catalog number 340450) and IL-4-PECy7 (clone 8D4-8, BD Pharmingen, catalog number 560672). Finally, stained cells were washed twice with Perm/Wash 1X and fixed in formaldehyde 1%. Cytokine responses were background subtracted. All samples were acquired on BD FACSCanto II (BD Biosciences) flow cytometer and analyzed using FlowJoTM v.10 software (Tree Star, Ashland, OR).

### Outcomes

Primary safety endpoints included the number and percentage of solicited local and systemic reactions through Day 7 after vaccination, unsolicited AEs through Day 28 after vaccination, grade 3 and 4 changes in safety laboratory parameters from baseline to Day 14, and SAEs related to study vaccine through end of the study. Secondary immunogenicity endpoints included neutralization titer against SARS-CoV-2 Wuhan,Beta, Delta and Omicron BA.1 variants measured as IC_50_ and reported as reciprocal concentration for each individual sample, GMT for descriptive statistics analysis at Baseline, Day 14, Day 91, Day 182 and Day 365, and GMFR from Baseline to Day 14, Day 91, Day 182 or Day 365. Secondary immunogenicity endpoints were also the binding antibody titers measured for each individual sample and GMT for descriptive statistics analysis at Baseline, Day 14, Day 91, Day 182 and Day 365.

Exploratory safety endpoints included the incidence of COVID-19 and the number and percentage of severe COVID-19 cases, both occuring from day 14 after administration of the booster and through end of the study. Number and percentage of hospital admissions, intensive care unit (ICU) admissions and non-invasive ventilation administration associated with COVID-19 were also reported from Day 14 to the end of the study. Exploratory immunogenicity endpoints included IFN-γ^+^ T-cell and CD4^+^/CD8^+^ T-cell responses to the SARS-CoV-2 S protein measured in re-stimulated PBMCs by ELISpot and ICS, respectively, at Baseline and Day 14.

### Statistical analysis

No formal sample size calculation was performed for this Phase III study. The following analyses of populations were included in the study: Intention-to-treat (ITT) population, including all subjects who were enrolled, regardless of the subject’s treatment status in the study; modified ITT (mITT) population, comprising all subjects in the ITT who met the eligibility criteria, received a dose of the vaccine and had not tested positive for COVID-19 within 14 days of receiving study drug; immunogenicity (IGP) population, comprising all subjects in the mITT who had a valid immunogenicity test result before receiving study drug and at least one valid result after dosing; and safety (SP) population comprising all enrolled subjects who received the study drug, and were analyzed according to their primary vaccination schemes.

Tabulations were produced for appropriate demographic, baseline, safety, and immunogenicity parameters. For categorical variables, summary tabulations of the number and percentage of subjects within each category of the parameter were presented. For continuous variables, the number of subjects, mean, standard deviation (SD), minimum and maximum values were presented, where appropriate. For the immunogenicity variables, the geometric mean and geometric standard deviations were presented, as appropriate.

Screening demographic characteristics (age, age category, sex, ethnicity, race, height [cm], weight [kg], and body mass index [BMI]), baseline vital signs (systolic and diastolic blood pressures [mmHg], pulse rate [beats per minute], pulse oximetry [%], and body temperature [°C]), primary COVID-19 vaccination group, time elapsed since last dose of primary COVID-19 vaccine (months), and prior COVID-19 infections were presented using summary statistics. No statistical comparisons were performed for any of the baseline characteristics.

A mixed effects model for repeated measures (MMRM) were carried out on log transformed data to measure the neutralization titer against Wuhan strain and beta, delta and omicron variants as measured by IC_50_ with PBNA. The weighted Least Squared (LS) mean estimates are presented with the associated standard error and 95% CIs. The back-transformed treatment group LS mean estimate for weighted LS means ratio (GMT) are presented with the corresponding 95% CI. Summary statistics for the log10 transformations for each individual sample were calculated based on the log10-transformed titers at Baseline, Day 14, Day 91, Day 182 and Day 365; and are presented for the Immunogenicity population. To evaluate the immunogenicity, total antibody binding antibodies titer measured for each individual sample and GMT for descriptive statistics analysis at Baseline and Days 14, 91, 182, and 365 were analyzed in a similar manner as described above, GMFR in neutralizing and binding antibodies were assessed using an analysis of variance model (ANOVA model) carried out on log transformed data. For the cellular immunogenicity analysis, a MMRM of T-cell data was employed, using the angular-transformed proportions as the response variable.

Analyses of AEs were performed for those events that were considered treatment-emergent, where treatment-emergent was defined as any AE with onset on or after the administration of study treatment until 28 days thereafter. TEAEs were presented by maximum intensity (Grade 1, 2, 3, and 4) and causal relationship to study drug (pooled as related or not related categories). Solicited local and systemic adverse reactions and unsolicited adverse events after dosing were also presented by maximum intensity and cumulatively across severity levels.

COVID-19 infections and other exploratory safety data are shown as the number of events and percentage of participants affected in the safety population. An exact 95% Clopper-Pearson CI for the proportion of each endpoint was also presented.

All descriptive statistical analyses were performed using SAS statistical software Version 9.4, unless otherwise noted. Medical history and adverse events were coded using MedDRA Version 26.0 and listed by SOC and PT.

## RESULTS

### Study participants

Overall, 2661 subjects were screened for this study and were boosted with PHH-1V vaccine. A flow diagram of the study participants is depicted in **Figure 1**. A total of 158 (5.94%) subjects prematurely discontinued participation in the study. The reasons for subject premature discontinuation included death (traffic accident), lost to follow-up, withdrawal by subject, and others. All the subjects enrolled in the study were included in the ITT and safety populations. A total of 2571 subjects were included in the mITT and 235 subjects were included in the IGP population for immunogenicity analysis. The mean study duration for subjects was 6.4 months (range: 0.03-12.65 months).

**Figure 1.**
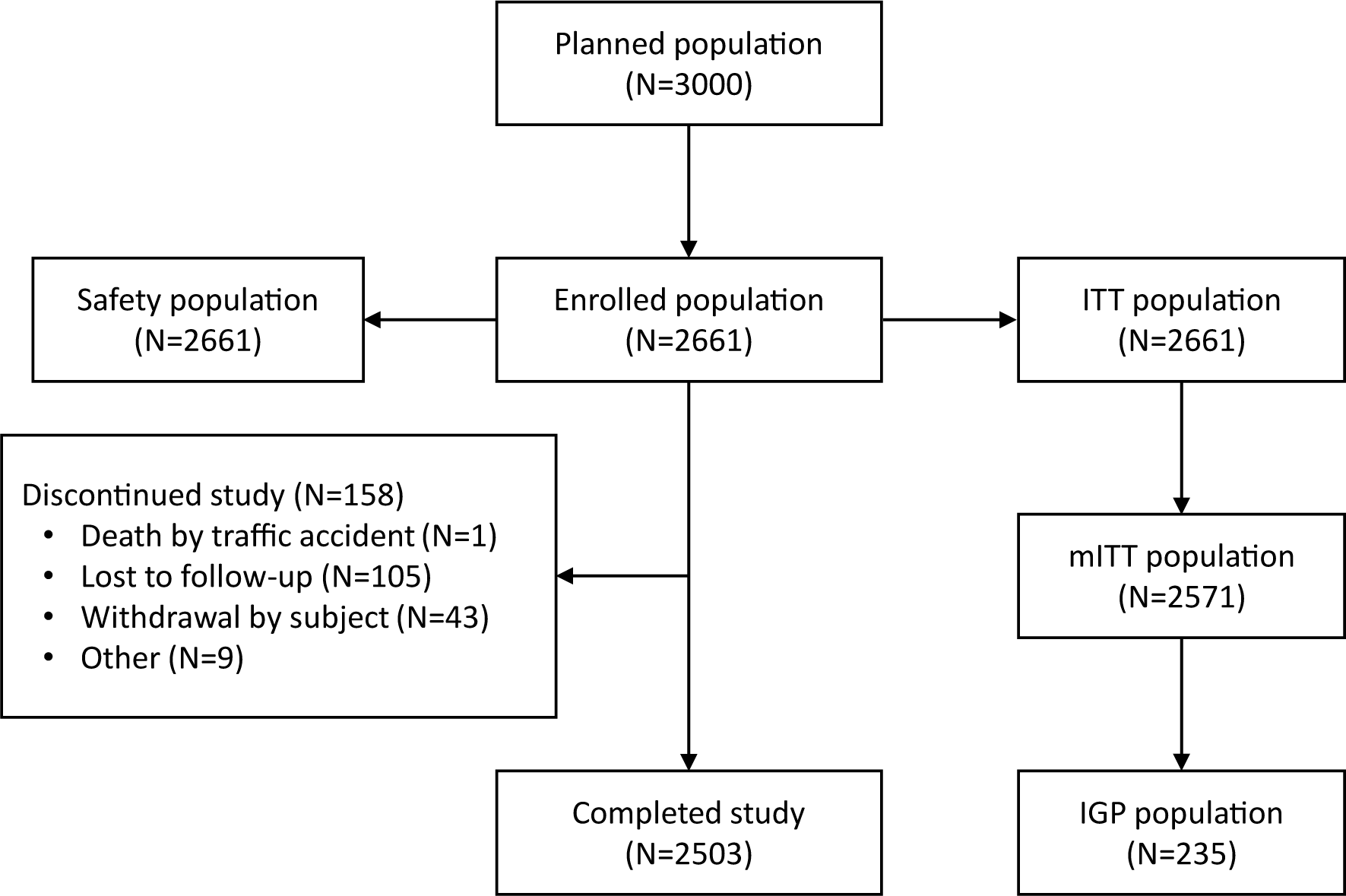
Flow chart of the study. ***Abbreviations:*** *IGP, immunogenicity population; ITT, Intention-to-treat; mITT, modified intention-to-treat; N, number of subjects meeting the criterion*.

A summary of demographics and baseline characteristics for the safety population is provided in **Table 1**. Overall, the mean age of participants was 34.5 years (range:16-85 years), 2603 (97.86%) were aged between 18 to 64 years, 1388 (52.16%) were male, 2633 (98.95%) were white, and 2242 (84.25%) were not Hispanic or Latino. The mean BMI was 24.69 kg/m^2^ (range: 14.86-51.90 kg/m^2^). At baseline, 1156 (43.44%) subjects had COVID-19 after their primary COVID-19 vaccination, of which 707 (26.57%) experienced COVID-19 ≤ 3 months after their primary vaccination, 66 (2.48%) experienced COVID-19 ˃ 3 months after their primary vaccination, and 377 (14.17%) experienced COVID-19 prior to their last primary vaccination. Of the subjects with prior COVID-19 infections, 695 (26.12%) were tested using RAT and 477 (17.93%) were tested using PCR. All infections prior PHH-1V vaccination were non-severe. Additionally, 1550 (58.25%) subjects received BNT162b2/ BNT162b2 as their primary COVID-19 vaccination, 565 (21.23%) received mRNA-1273/ mRNA-1273, and 112 (4.21%) received ChAdOx1-S/ ChAdOx1-S. Most of the subjects (70.12%) received PHH-1V booster > 6 to ≤ 12 months after their primary COVID-19 vaccination.

**Table 1.** Demographic profile and baseline characteristics of participants. Demographics and baseline characteristics were described for the safety population. Percentages were calculated as (%) = n/N*100. *Subjects with one dose of COVID-19 vaccination and COVID-19 infection (before or after the vaccination). ***Abbreviations:*** *COVID-19, corona virus disease 2019; N, the number of subjects in the population; n, the number of subjects meeting the criterion; SD, standard deviation*.

| Characteristics | PHH-1V (N=2661) |
| --- | --- |
| **Age (years)** |  |
| Mean (SD) | 34.5 (12.79) |
| Min-Max | 16-85 |
| **Age group, n (%)** |  |
| ≥ 16 to < 18 years | 36 (1.35) |
| ≥ 18 to < 65 years | 2603 (97.86) |
| ≥ 65 years | 22 (0.79) |
| **Sex, n (%)** |  |
| Male | 1388 (52.16) |
| Female | 1272 (47.80) |
| Undifferentiated | 1 (0.04) |
| **Race, n (%)** |  |
| White | 2633 (98.95) |
| Black or African American | 8 (0.30) |
| American Indian or Alaska Native | 7 (0.26) |
| Asian | 2 (0.08) |
| Other | 11 (0.41) |
| **Previous COVID-19 infection, n (%)** |  |
| ≤ 3 months after primary vaccination | 707 (26.57) |
| > 3 months after primary vaccination | 66 (2.48) |
| Prior to the last primary vaccination | 377 (14.17) |
| Missing vaccination date | 2 (0.08) |
| Other missing | 4 (0.15) |
| No | 1505 (56.56) |
| **Primary COVID-19 vaccination group, n (%)** |  |
| BNT162b2* | 197 (7.40) |
| BNT162b2/ BNT162b2 | 1550 (58.25) |
| BNT162b2/ mRNA-1273 | 13 (0.49) |
| Ad26.COVS2-S* | 48 (1.80) |
| Ad26.COVS2-S / BNT162b2 | 19 (0.71) |
| Ad26.COVS2-S / Ad26.COVS2-S | 7 (0.26) |
| Ad26.COVS2-S / mRNA-1273 | 12 (0.45) |
| mRNA-1273* | 90 (3.38) |
| mRNA-1273/ mRNA-1273 | 565 (21.23) |
| mRNA-1273/ ChAdOx1-S | 2 (0.08) |
| ChAdOx1-S* | 5 (0.19) |
| ChAdOx1-S/ BNT162b2 | 37 (1.39) |
| ChAdOx1-S/ mRNA-1273 | 1 (0.04) |
| ChAdOx1-S/ ChAdOx1-S | 112 (4.21) |
| Others | 3 (0.11) |
| **Time elapsed since last primary COVID-19 vaccination dose or infection, n (%)** |  |
| ≤ 3 months | 26 (0.98) |
| > 3 to ≤ 6 months | 721 (27.10) |
| > 6 to ≤ 12 months | 1866 (70.12) |
| > 12 months | 45 (1.69) |
| Other missing | 3 (0.11) |

### Safety and tolerability

The safety analysis included 2661 subjects who received a dose of the study vaccine. TEAE incidence is summarized in **Table 2**. A total of 7573 TEAEs were reported in 2347 (88.20%) subjects. Most TEAEs were Grade 1 (58.70%) or Grade 2 (27.58%) intensity. Overall, 22 (0.83%) subjects had TEAEs not related to the administration of PHH-1V and 2325 (87.37%) subjects had TEAEs related to the administration of PHH-1V. TEAE reported in ≥1.0% of subjects are summarized in **Table S3**. The most common TEAEs were injection site pain (82.83%), fatigue (31.72%), headache (31.23%) and myalgia (20.74%).

**Table 2.** Summary of Treatment-Emergent Adverse Events (TEAEs). TEAEs were described for the safety population (N=2661). A TEAE was defined as an AE that started on or after the date of administration of study treatment until 28 days thereafter. If AE dates were incomplete and it was not clear whether the AE was treatment-emergent, it was assumed to be treatment-emergent. ^1^If a subject experienced more than one TEAE, It is counted once at the most severe or most related event. 2Unrelated adverse events are those classified as not related and unlikely related. 3Related adverse events are those classified as possibly, probably and related. If an AE has a missing relationship it is assumed to be related to the study treatment for analysis purposes. *Solicited AEs reported up to day 7. **Unsolicited AEs up to day 28. ‡SAEs up to study end. †Car crash ***Abbreviations****: AE,* adverse *event; SAE, serious adverse event; N, the number of subjects in the population; TEAE, treatment-emergent adverse event*.

|  | PHH-1V (N=2661) |  |
| --- | --- | --- |
|  | Events | Subjects (%) |
| **Total number of TEAEs** | 7573 | 2347 (88.20) |
| **TEAE intensity<sup>1</sup>** |  |  |
| Grade 1 | 6173 | 1562 (58.70) |
| Grade 2 | 1329 | 734 (27.58) |
| Grade 3 | 68 | 49 (1.84) |
| Grade 4 | 3 | 2 (0.08) |
| **TEAE relationship to study treatment** |  |  |
| Unrelated<sup>2</sup> | 434 | 22 (0.83) |
| Related<sup>3</sup> | 7139 | 2325 (87.37) |
| **Total number of TEAEs leading to death<sup>†</sup>** | 1 | 1 (0.04) |
| **Total number of solicited local and systemic AEs\*** | 6861 | 2320 (87.19) |
| **Intensity of solicited local and systemic AEs<sup>1</sup>** |  |  |
| Grade 1 | 5577 | 1583 (59.49) |
| Grade 2 | 1226 | 695 (26.12) |
| Grade 3 | 56 | 41 (1.54) |
| Grade 4 | 2 | 1 (0.04) |
| **Total number of unsolicited AE\*\*** | 712 | 538 (20.22) |
| **Intensity of unsolicited adverse events<sup>1</sup>** |  |  |
| Grade 1 | 596 | 437 (16.42) |
| Grade 2 | 103 | 89 (3.34) |
| Grade 3 | 12 | 11 (0.41) |
| Grade 4 | 1 | 1 (0.04) |
| **Total number of SAEs<sup>‡</sup>** | 22 | 21 (0.79) |
| **Intensity of SAEs<sup>1</sup>** |  |  |
| Grade 2 | 6 | 6 (0.23) |
| Grade 3 | 13 | 12 (0.45) |
| Grade 4 | 3 | 3 (0.11) |
| **Relationship of SAEs to study treatment** |  |  |
| Unrelated<sup>2</sup> | 21 | 20 (0.75) |
| Related<sup>3</sup> | 1 | 1 (0.04) |

A total of 6861 solicited local and systemic adverse events were reported in 2320 (87.19%) subjects, which were mostly Grade 1 (59.49%) or Grade 2 (26.12%) intensity (**Table 2**). Of these, 6857 solicited local reactions and systemic events were related to the administration of PHH-1V. The most reported number of solicited local reactions was on Day 0 with 3498 events in 1912 (71.85%) subjects, decreasing each day through to Day 7 (1.92% of subjects) (**Figure 2A**). The most frequently reported solicited local reactions from Day 0 to Day 7 were pain and tenderness, with 64.37% of subjects experiencing pain and 57.57% of subjects experiencing tenderness on Day 0 and decreasing to 1.28% and 1.43%, respectively, on Day 7. On Day 0, the number of solicited systemic events was 1285 in 748 (28.11%) subjects, which peaked on Day 1 (32.69%) and then decreased thought to Day 7 (5.79%) (**Figure 2B**). The most common solicited systemic events from Day 0 through Day 7 were fatigue, headache and muscle pain, with 19.92% of subjects experiencing fatigue, 17.47% experiencing headache and 13.45% experiencing muscle pain on Day 1, and decreasing to 2.97%, 2.93% and 1.99%, respectively, on Day 7.

**Figure 2.**
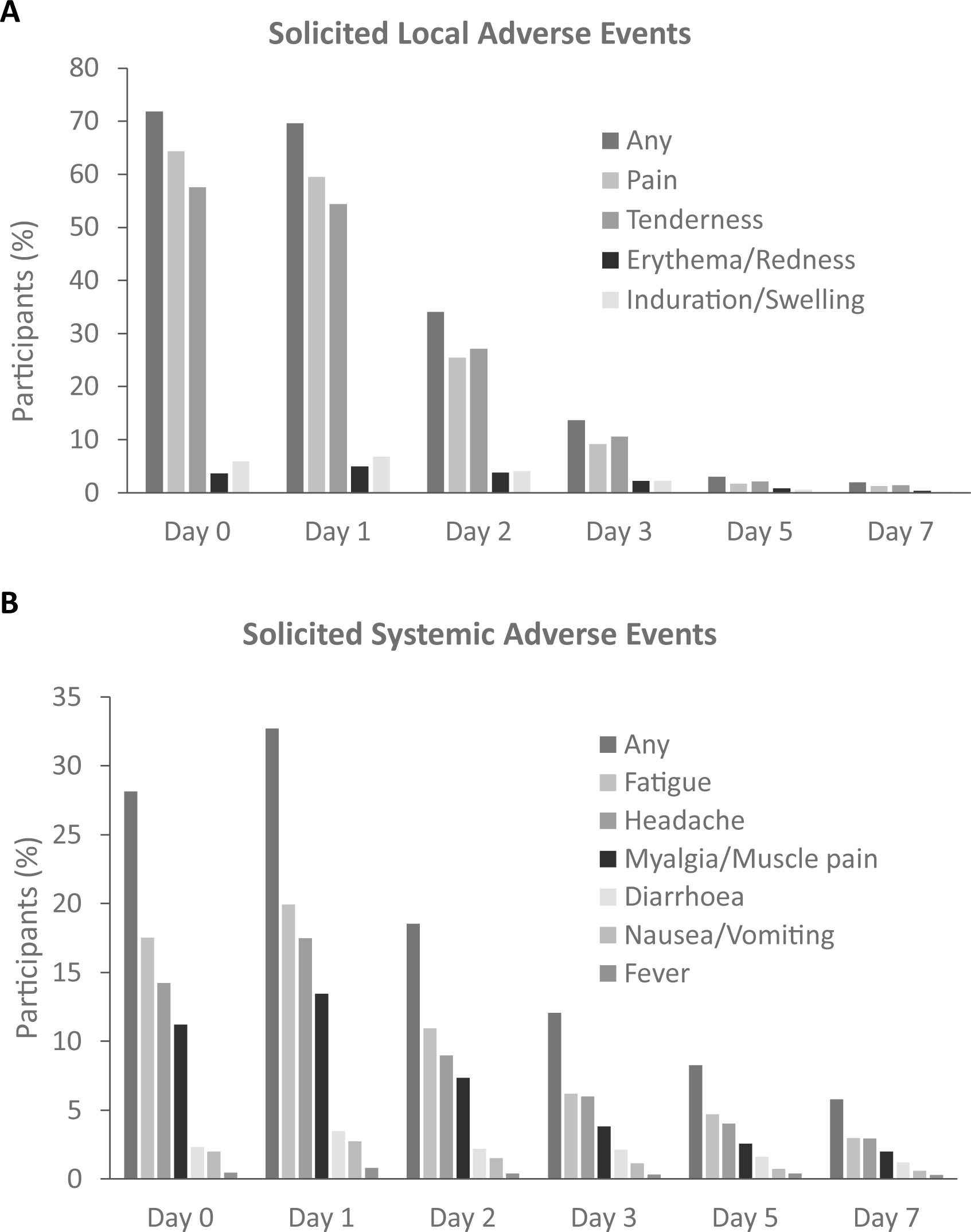
Solicited local and systemic adverse events through Day 7. Solicited local adverse events (A) and solicited systemic adverse events were reported by MedDRA Preferred Term (PT) from Day 0 through Day 7 for the safety population. Data are shown as the percentage of subjects in relation to the safety population (N=2661). If a subject experienced more than one event, the subject is counted once for each type of event. PTs are ordered in decreasing frequency of the total number of subjects with each adverse event. ***Abbreviations:*** *N, the number of subjects in the population; PT, preferred term*.

Unsolicited AEs were reported in 538 (20.22%) of subjects, most of which were Grade 1 intensity (16.42% of subjects) (**Table 2**). In total, 430 unsolicited AEs were not related to the administration of PHH-1V in 311 (11.69%) subjects and 282 were related to the administration of PHH-1V in 227 (8.53%) subjects. Unsolicited adverse events from Day 0 through to Day 28 in ≥1% of overall subjects are summarized in **Table S4**. The most frequently reported unsolicited AE up to day 28 was COVID 19 infection with 53 events in 53 (1.99%) subjects. Other unsolicited AEs were axillary pain (1.39%), lymphadenopathy (1.24%) and injection site induration (1.16%).

Over the entire study period, 22 SAEs were reported in 21 (0.79%) subjects (**Table 2**). The most frequently reported SAEs were joint dislocation (0.08%) and appendicitis (0.08%), but no SAEs were reported above 1% of the overall population. One SAE, a Grade 2 pericarditis on a male between 35-40 years old, was considered related due to temporal relationship to the administration of PHH-1V and was reported as a SUSAR. The event appeared 14 days after PHH-1V administration and was considered resolved 143 days after. A road traffic accident was reported as fatal SAE unrelated to the administration of PHH-1V.

Vital signs measurements (systolic and diastolic blood pressure, pulse rate, oxygen saturation, or temperature) were within normal clinical values, and no significant changes occurred in hematology or biochemistry laboratory evaluations among the subjects during the period of this study report.

No relevant differences in the safety profile were observed between different primary vaccination schedules, and none of the previous primary vaccinations showed more reactogenicity than others after administration of the PHH-1V booster dose (data not shown). Similarly, subjects with a previous history of COVID-19 had no increase in reactogenicity when receiving PHH-1V independently of the time elapsed from this previous infection.

### Neutralizing and binding antibody responses

Immunogenicity was assessed at baseline, Day 14, Day 91, Day 182, and Day 365 in a subset of 235 subjects vaccinated with two doses of BNT162b2/BNT162b2 (individuals 16-17 years old; n=13), mRNA-1273/mRNA-1273 (n=172) or ChAdOx1-S/ChAdOx1-S (n=42), or a combination of ChAdOx1-S and another brand of vaccine (n=8, 7 were vaccinated with ChAdOx1-S/BNT162b2 and one with mRNA-1273/ChAdOx1-S). The GMT of neutralizing antibodies determined by PBNA at baseline, Day 14, Day 91, Day 182 and Day 365 post-booster for overall participants independent of prime vaccination group and GMFR against baseline at day 14, day 91, day 182 and day 365 post-boost are shown in **Table 3** and represented on **Figure 3**. To avoid potential bias in immunogenicity assessment, all subjects who reported SARS-CoV-2 infection were excluded from the neutralizing and binding antibody analysis. Overall, GMT for neutralizing antibodies increased on Day 14 for all SARS-CoV-2 variants analyzed with a significant increase compared to baseline titers with GMFR of 6,90 (95% CI 4,96-9,58) for Wuhan, 12,27 (95% CI 8,52-17,67) for Beta, 7,24 (95% CI 5,06-10,37) for Delta and 17,51 (95% CI 12,28-24,97) for Omicron BA.1. A subsequent decrease on GMT for adjusted treatment over time was observed for all analyzed variants but, in all cases, remained significatively different from baseline over all time points up to 1 year after PHH-1V booster administration (**Figure 3A** and **Table 3**). GMT for adjusted treatment with its 95% CI for different prime vaccination groups are shown on **Figure 3B** and **Table S5** against Wuhan, Beta, Delta and Omicron BA.1 variants. Booster dose with PHH-1V drives a significative increase on neutralized antibodies at day 14 against all SARS-CoV-2 variants analyzed regardless of the prime vaccination scheme (**Table S7**). All prime vaccination groups show a decrease on GMT for adjusted treatment after its induction at day 14, but still numerically higher than baseline for all variants and prime vaccination groups up to 1 year after booster vaccination with PHH-1V, except for group vaccinated with ChAdOx1-S and another brand of vaccine (n=8, 7 were vaccinated with ChAdOx1-S/BNT162b2 and one with mRNA-1273/ChAdOx1-S), these 8 participants show a sustained and significative levels of neutralizing antibodies titers up to 1 year after booster with PHH-1V compared to baseline for all SARS-CoV-2 variants (**Figure 3B** and **Table S5)**.

**Figure 3:**
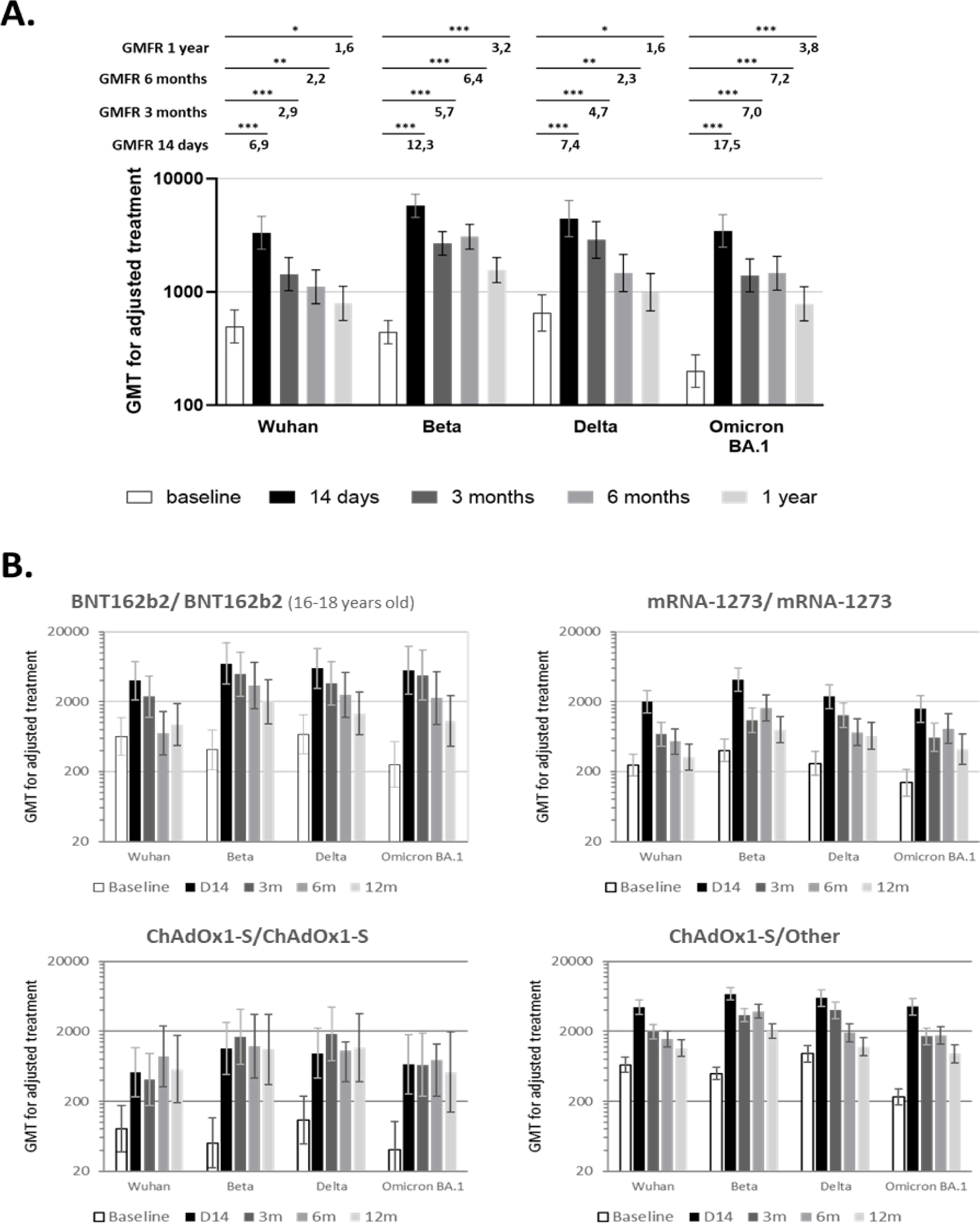
Neutralizing antibody levels against SARS-CoV-2 variants after booster with PHH-1V over time. (A.) Representation of Mean GMT for adjusted treatment with the 95% CI (graphics) and mean GMFR (Upper numbers) from baseline against SARS-CoV-2 Wuhan, Beta, Delta and Omicron BA.1 variants in overall subjects (n= 235) at baseline (White) and Days 14 (Black), 3 months (Dark grey), 6 months (Grey) and 1 year (Light grey) post-boost. *** p< 0.0001; ** p< 0.001; * p<0,01. (B.) Mean GMT for adjusted treatment with the 95% CI over time for each prime vaccination group. BNT162b2/ BNT162b2 (n=13); mRNA-1273/ mRNA-1273 (n=172); ChAdOx1-S/ChAdOx1-S (n=42) and ChAdOx1-S/other (n=8). *Subjects who reported COVID-19 infections were excluded from the reported day onwards*. ***Abbreviations :*** *CI = confidence interval; GMT = Geometric Mean Titer; GMFR = Geometric Mean Fold Rise*.

**Table 3.**
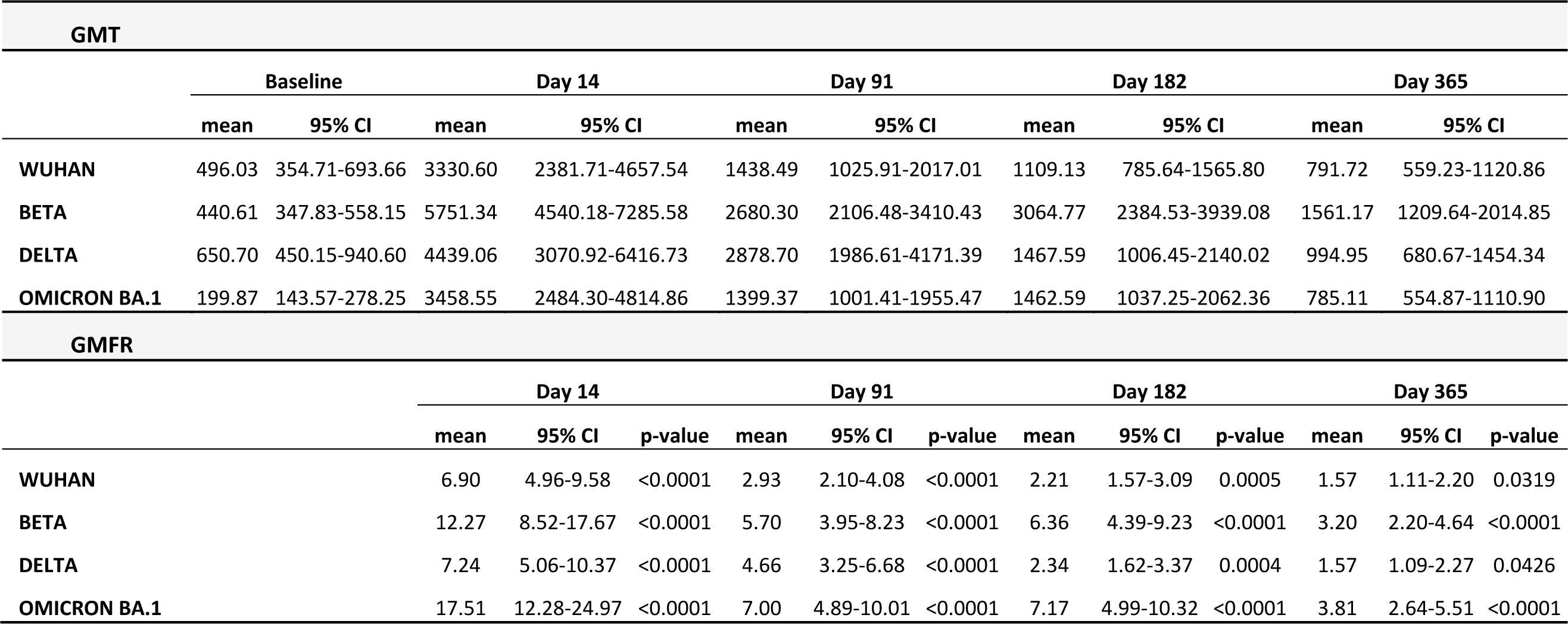
Analysis of neutralizing antibody against SARS-CoV-2 variants after a booster dose with PHH-1. Mean GMTs for adjusted treatment and corresponding 95% CI against SARS-CoV-2 Wuhan, Beta, Delta and Omicron BA.1 variants in overall subjects (n= 235) at baseline and Days 14, 91, 182 and 365 post-boost are shown. Mean GMFR calculated between time point GMT for adjusted treatment and baseline GMT with GMFR p-value for odds ratio=1 were shown. Raw data provided as < 20 have been imputed as 20 for the purposes of analysis. Any zero values have been imputed to 10 (half the LLOQ) for analysis purposes. In all timepoints except Day 365/ETV, raw data provided as > 20480 have been imputed as 20480 for the purposes of analysis. *Subjects who reported COVID-19 infections were excluded of this analysis from the report day onwards*. ***Abbreviations:*** *CI, confidence interval; GMT, geometric mean titer; GMFR, geometric mean fold rise; ETV, early termination visit; LLOQ, lower limit of quantification*.

Binding antibody responses are summarized in **Table 4**. Overall, a high binding antibody response was observed on Day 14 after the booster in all primary vaccination groups. The GMT (95% CI) for binding antibodies against SARS-CoV-2 RBD on Day 14 were 63866.67 (35790.22, 113968.32) for BNT162b2/BNT162b2, 51520.75 (41401.54, 64113.25) for mRNA-1273/mRNA-1273, 18443.18 (13149.13. 25868.71) for ChAdOx1-S/ChAdOx1-S and 13666.18 (6597.12, 28309.99) for ChAdOx1-S/Another brand. Furthermore, GMFR (95% CI) for binding antibodies against SARS-CoV-2 on Day 14 were 12.98 (5.48, 30.77) for BNT162b2/BNT162b2, 16.82 (10.66, 26.53) for mRNA-1273/mRNA-1273, 14.82 (8.38, 26.23) for ChAdOx1-S/ChAdOx1-S and 30.22 (10.51, 86.90) for ChAdOx1-S/Another brand (**Table 4**). GMTs for binding antibodies gradually decline on Day 91, Day 182 and Day 365 but are still higher than baseline titers. On Day 365, GMT (95% CI) against SARS-CoV-2 were 9390.52 (5085.21, 17340.85), 4257.94(2940.99, 6164.62), 6388 (2493.64, 16364.29) and 10404.04 (8221.36, 13166.21) for the BNT162b2/BNT162b2, ChAdOx1-S / ChAdOx1-S, ChAdOx1-S/Another, and Spikevax/Spikevax vaccine groups, respectively; and GMFR (95% CI) were 1.66 (0.66, 4.20), 2.88 (1.54, 5.37), 14.8 (3.51, 62.53) and 2.88 (1.79, 4.65) for the BNT162b2/BNT162b2, ChAdOx1-S / ChAdOx1-S, ChAdOx1-S/Another, and mRNA-1273/mRNA-1273 vaccine groups, respectively.

**Table 4.** Analysis of binding antibody against SARS-CoV-2 after a booster dose of PHH-1V. Binding antibodies titers were analyzed against SARS-CoV-2 in a subset of subjects primary vaccinated with BNT162b2/BNT162b2 (N=13), mRNA-1273/mRNA-1273 (N=172), ChAdOx1-S/ChAdOx1-S (N=42) and, ChAdOx1-s/Another (N=8) at baseline and Days 14, 91, 182 and 364 post-boost. Binding antibodies against the SARS-CoV-2 RBD was assessed by the percentage of subjects having a ≥4-fold increase in the binding antibodies titer 14, 91, 182, and 365 days after booster using the Elecsys Anti-SARS-Cov-2 S immunoassay. A MMRM model was fitted to assess the endpoint on the log10 scale. GMTs, GMFRs and the corresponding 95% Cl were estimated using LS Means from MMRM model fitted on the log10 scale and then back-transformed. *p<0.05; **p<0.001; ***p<0.0001. Raw data provided as > 225000 have been imputed as 225000 for the purposes of analysis. *Subjects who reported COVID-19 infections were excluded of this analysis from the report day onwards*. ***Abbreviations****: CI, confidence interval; GMFR, geometric mean fold rise; GMT, geometric mean titer; LS mean, least square mean; MMRM, mixed effects model for repeated measures ;N, number of subjects in each primary vaccination group; RBD, receptor binding domain*.

|  | BNT162b2/BNT162b2<br>(N=13) | mRNA-1273/mRNA-1273<br>(N=172) | ChAdOx1-S/ChAdOx1-S<br>(N=42) | ChAdOx1-S/ Another<br>(N=8) |
| --- | --- | --- | --- | --- |
| **GMT [95% CI]** |  |  |  |  |
| Baseline | 4268.34 [2391.94 - 7616.74] | 3426.17 [2753.24 - 4263.59] | 1079.44 [731.10 - 1678.54] | 490.52 [236.79 - 1016.14] |
| Day 14 | 63866.67 [35790.22 - 113968.32] | 51520.75 [41401.54 - 64113.25] | 18443.18 [13149.13 - 25868.71] | 13666.18 [6597.12 - 28309.99] |
| Day 91 | 31183.96 [17215.09 - 56487.63] | 27862.81 [22303.89 - 34807.21] | 8631.33 [6084.78 - 12243.64] | 11737.91 [5466.09 - 25206.04] |
| Day 182 | 14236.33 [7551.76 - 26837.86] | 17521.55 [13900.35 - 22086.12] | 5806.01 [4025.48 - 8374.09] | 11740.62 [4941.62 - 27894.14] |
| Day 365 | 9390.52 [5085.21 - 17340.85] | 10404.04 [8221.36 - 13166.21] | 4257.94 [2940.99 - 6164.62] | 6388 [2493.64 - 16364.29] |
| **GMFR [95% CI]** |  |  |  |  |
| Day 14 | 13.23 [5.77 - 30.34]*** | 15.57 [10.82 - 22.41]*** | 16.13 [9.77 - 26.60]*** | 30.25 [10.75 - 85.13]*** |
| Day 91 | 6.5 [2.82 - 14.97]*** | 8.61 [5.98 - 12.40]*** | 7.54 [4.56 - 12.49]*** | 27.7 [9.72 - 78.92]*** |
| Day 182 | 3.28 [1.41 - 7.63]* | 5.52 [3.82 - 7.96]*** | 5.14 [3.09 - 8.55]*** | 28.49 [9.70 - 83.69]*** |
| Day 365 | 2.00 [0.87 - 4.64] | 3.3 [2.28 - 4.76]*** | 3.76 [2.26 - 6.26]*** | 16.01 [5.33 - 48.10]*** |

### T-cell responses

The RBD/Spike-specific IFN-γ^+^ T-cell response was analyzed by ELISpot in a subset of 24 subjects immunized with PHH-1V after receiving a primary vaccination with ChAdOx1-S. Of these, 19 had valid ELISpot measurements at Baseline and Day 14 and were eligible for analysis. Immunization with the PHH-1V vaccine, after a primary vaccination with ChAdOx1-S, showed a significant activation of IFN-γ-producing cells after the *in vitro* re-stimulation with peptide pools of Spike SA (p=0.0019) and RBD (Wuhan, p=0.0005; alpha, p=0.0121; beta, p=0.0007; delta, p=0.0352) at 2 weeks post-boost (Day 14) compared to the activation observed at baseline (**Figure 4**). No significant differences were observed between IFN-γ-producing cells elicited at baseline and Day 14 when PBMC were stimulated with Spike SB peptide pool. As the Spike B peptide pool did not contain RBD sequences, it was not expected an increase in T-cells activation after vaccination.

**Figure 4.**
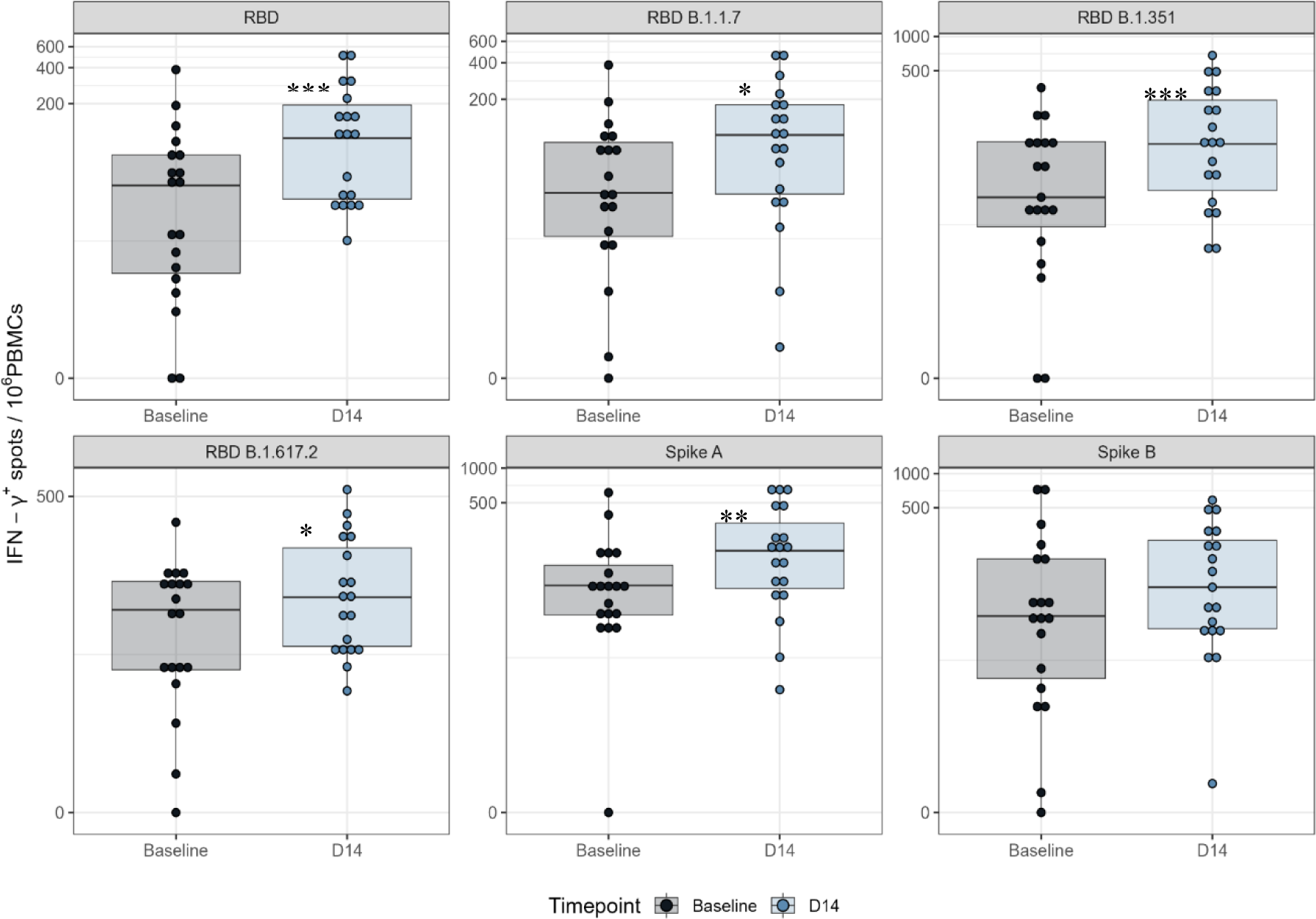
Total IFN-γ producing T cells assessed by ELISpot at baseline and Day 14. Frequencies of IFN-γ responses determined by ELISpot assay in PBMC from subjects immunized with PHH-1V after a primary vaccination with Vaxzevria. PBMC were isolated before the boost immunization (visit 2) and two weeks after boost with PHH-1V (visit 3), stimulated with RBD (Wuhan, RBD alpha, RBD beta and RBD delta) and Spike (SA, SB) peptide pools, and analyzed by IFN-γ-specific ELISpot assay. T-cell responses are expressed as IFN-γ^+^ spots per 10^6^ PBMCs. Boxes depict the median (solid line) and the interquartile range (IQR), and whiskers expand each box edge 1.5 times the IQR. A logarithmic scale has been used for plotting purposes. *p< 0,05; **p< 0,01; ***p<0,001 ***Abbreviations:*** *IFN-γ, interferon gamma; IQR, interquartile range; PBMC, peripheral blood mononuclear cell; RDB; receptor binding domain for the SARS-CoV-2 spike protein (ancestor Wuhan-Hu-1 strain); RDB B.1.1.7 (alpha variant); RDB B.1.351 (beta variant); Spike SA corresponds to 194 spike protein peptide pools overlapping the S1-2016 to S1-2196 region of the Spike protein; Spike SB corresponds to 168 spike protein peptide pools overlapping the S1-2197 to S2-2377 region of the Spike protein*.

Furthermore, the frequency of CD4^+^ and CD8^+^ T-cells expressing IFN-γ, IL-2, and IL-4 was assessed by ICS at two weeks post-boost (Day 14) in the same subset of 24 subjects. Of these, 15 had valid ICS measurements at Baseline and Day 14 and were eligible for analysis. Immunization with PHH-1V, after a primary vaccination with ChAdOx1-S, elicited CD4^+^ and CD8^+^ T cell expressing cytokines upon the stimulation with the RBD (Wuhan strain or alpha, beta or delta variants) and Spike (SA or SB) peptides on Day 14 post-boost (**Figure S1 and S2**). In particular, PHH-1V vaccination elicited significantly higher CD4^+^ T cell levels expressing IFN-γ upon stimulation with Wuhan (p=0.0073), alpha (p=0.0191) and beta (p=0.0115) peptide pools; and higher CD4^+^ T cell levels expressing IL-4 upon stimulation with beta (p=0.0331) and SA (0.0229) peptide pools compared to baseline levels. No significant increases were observed in the activation of CD8^+^ T-cells (Figure S2).

### Prevalence of COVID-19 in vaccinated population

No subjects experienced severe COVID-19, none were hospitalized due to COVID-19, none required non-invasive ventilation administration, none were admitted to the ICU due to COVID-19 ≥ 14 days after PHH-1V booster and throughout the study duration. There had been no deaths associated with COVID-19 after PHH-1V booster and throughout the study duration. Overall, 43 subjects (1.62% [95% CI: 1.17, 2.17]) had a SARS-CoV-2 infection < 14 days after PHH-1V booster, and 397 non-severe COVID-19 cases in 397 (14.92% [95% CI: 13.59, 16.33]) subjects were reported ≥ 14 days after PHH-1V booster and throughout the study duration.

## DISCUSSION

Herein we present study results from a Phase III, open-label, single arm, multicenter clinical study in healthy adults fully vaccinated against COVID-19. The primary objective was to assess the safety and tolerability of PHH-1V as a booster dose in subjects previously vaccinated against COVID-19 with the BNT162b2, ChAdOx1-S, mRNA-1273 or Ad26.COV2-S vaccines including those that had been previously infected by SARS-CoV-2. At present, many of the vaccines presented on this trial results has been adapted following recommendations, although the novelty resides on results showing that a booster dose of PHH-1V vaccine is immunogenic for up to 1 year and safe in individuals primary vaccinated with any of the COVID-19 vaccination scheme recognized by the European authorities at the study period.

This study included 2661 participants aged 16-85 years (mean age: 34.5 years), with a predominance of non-Hispanic and non-Latino white subjects. Most of the subjects did not experience a previous COVID-19 infection, and the rest of the participants reported non-severe COVID-19. The most common primary vaccination series were BNT162b2/ BNT162b2 and mRNA-1273/ mRNA-1273, and most subjects received their full primary vaccination schedule 6-12 months ago. Our study population mimiqued the EU situation at the time of study period, since the most frequently received vaccine in the European Union are BNT162b2, followed by mRNA-1273 and ChAdOx1-S^26^.

Frequency of adverse events reported after a booster dose with PHH-1V (88.20 %) were similar or even lower than the frequencies reported in the phase IIb trial^21^. Reactogenicity was assessed for 7 days after the booster dose with PHH-1V vaccine. PHH-1V was well-tolerated and had a low reactogenicity in all the primary vaccination groups, with most of the solicited adverse reaction being grade 1. The most common solicited local adverse reactions were pain and tenderness, and the most common solicited systemic adverse reactions were fatigue and headache. The safety profile of PHH-1V described in this phase III clinical trial was very similar to the safety profile reported previously in the phase IIb trial^21^. Although the relative frequencies of local and systemic reactions are difficult to compare across studies, the reactogenicity profile of the vaccine was also consistent with those observed after a booster dose with subunit- and RNA-based vaccines^13,25,27,28^. Indeed, a recent clinical study evaluating the safety of a third dose with the protein subunit vaccines NVX-CoV2373 (Nuvaxovid, Novavax) and SII NVX-CoV2373 (Covovax; Indian Serum Institute) showed that the most common solicited local adverse reactions were pain and tenderness, and the most common solicited systemic adverse reaction were headache and fatigue^29^. No relevant differences in the safety profile of PHH-1V were observed between different primary vaccination schedules. Furthermore, unsolicited adverse events were reported from Day 0 through Day 28 of the study, and SAE events were collected through the end of the study. Frequency of unsolicited AEs was low (20.22%) with most of the unsolicited AEs being grade 1. Similarly, frequency of SAE was low regardless of primary vaccination and most of them were unrelated with the PHH-1V administration. One grade 2 pericarditis was reported and considered the only related SAE due to temporal relationship to the administration of PHH-1V. Pericarditis was reported as a very rare AE in COVID-19 mRNA vaccinations, especially in young adult and adolescent males. In addition, subjects with a previous history of COVID-19 had no increase in reactogenicity when receiving PHH-1V independently of the time elapsed from this previous infection. This contrasts with a previous ChAdOx1-S (Vaxzevria; AstraZeneca), Ad26.COV2-S (Jcovden; Janssen) and mRNA-1273 (Spikevax; Moderna) booster vaccines based on the primary vaccination group^15^.

Vaccination with a booster dose of a COVID-19 vaccine has been associated with decreased risk of developing COVID-19-related symptoms, hospitalization, and death^5,31^. Another study in the US reported a significant increase of effectiveness in preventing COVID-19 associated hospitalizations with a primary COVID-19 vaccine schedule plus booster doses compared to a primary vaccine schedule alone^32^. Our results are in line with these findings since no subjects died due to COVID-19, none experienced severe COVID-19, none were hospitalized or required non-invasive ventilation administration due to COVID-19 or were admitted to the ICU due to COVID-19 ≥ 14 days after PHH-1V booster and throughout the study duration. Only 14.92% of subjects reported non-severe COVID-19 cases ≥ 14 days after PHH-1V booster and throughout the study duration.

Neutralizing antibodies are accepted correlates of COVID-19 vaccine efficacy since vaccines inducing high neutralizing antibodies titers (such as NVX-CoV2373, mRNA-1273 and BNT162b2) have shown higher vaccine efficacy in clinical trials than those associated with lower titres^33,34^. In our study, neutralizing antibodies levels were assessed at Baseline, Day 14, Day 91, Day 182, and Day 365 in a subset of 235 subjects without COVID-19 reported and vaccinated with two doses of BNT162b2/ BNT162b2 (individuals with 16-17 years), mRNA-1273/mRNA-1273, ChAdOx1-S/ChAdOx1-S, or a combination of ChAdOx1-S and another brand of vaccine. Overall, PHH-1V vaccine was able to elicit high increase of neutralizing antibody titers 14 days after the booster vaccination, against the Wuhan, Beta, Delta, and Omicron BA.1 SARS-CoV-2 variants compared to those levels at baseline, regardless of the primary vaccination schedule. GMTs for neutralizing antibodies against SARS-CoV-2 gradually dropped on days 91, 182 and 365 of the study, but were still significantly higher than the baseline titers. Only the small group of ChAdOx1-S with another brand of vaccine (n=8; 7 vaccinated with ChAdOx1-S/BNT162b2and one with mRNA-1273/ChAdOx1-S) has not shown this drop on neutralizing antibodies, this could be an effect of the small population included or due to unreported COVID-19 infection over study period. Remarkably, a booster vaccination with PHH-1V was able to sustain a good neutralizing activity against the Omicron BA.1 SARS-CoV-2, the most common variant at the time of starting this trial^35^. Compared to the Wuhan-Hu-1 reference genome, the RBD sequence of Omicron BA.1 comprises 15 mutations localized within the binding sites of epitopes targeted by monoclonal antibodies^36^. K417N substitution, present also in the Beta variant, is one of the 15 RBD substitutions in Omicron variant and responsible for a significant disruption to known monoclonal antibodies^37^. This fact strongly supports the high humoral immunogenicity of the PHH-1V RBD-based candidate against a wide range of potential new mutations since the PHH-1V antigen comprises key mutations that are also present in the Omicron BA.1 variant, as well as in many other SARS-CoV-2 variants. Of note, all subjects who reported SARS-CoV-2 infection were excluded from the neutralizing antibody analysis, thus eliminating the possible bias in the GMT measurement due to infection-induced immunogenicity.

A minimum clinically important difference of 1.75-times difference in GMT between pre- and post-booster levels has been proposed based on advice by UK policy makers^15^. Following this criteria, fold increase of neutralizing antibodies on Day 14 after the PHH-1V booster was clinically relevant regardless the primary vaccination series received with a GMFR of 6.90 against SARS-CoV-2 Wuhan, 12.27 against beta variant, 7.24 against delta variant and 17.51 against omicron BA.1 variant. Although the neutralizing antibody GMT decreased throughout the study, GMFR were still over 1.75 on Day 365 against beta (3.20; 95% CI: 2.20-4.64) and omicron BA.1 (3.81; 95% CI: 2.64-5.51) variants for most of the primary vaccination group showing that PHH-1V elicits a long term (one year) neutralizing response against a variety of SARS-CoV-2 strains including new emerging variants at that time point. In addition, binding antibodies were assessed at Baseline, Day 14, Day 91, Day 182, and Day 365 post-booster vaccination. In general, a high response in binding antibodies and fold rise in binding antibodies was observed 14 days after the PHH-1V booster dose. Even though a steady decrease in binding antibodies GMT were observed on Day 91, 182 and 365 after PHH-1V booster dose, titers were still higher compared to those at baseline. All together, these data demonstrate that a booster dose of PHH-1V can increase and maintain a humoral immune response over a long period of time.

While neutralizing antibodies play a primary role in preventing symptomatic COVID-19, the prevention of severe cases is likely influenced by various immune components such as CD4^+^ T cells, CD8^+^ T cells, and memory B cells^38–40^. Here, RBD/Spike-specific IFN-γ^+^ T-cell response was analyzed on Day 14 by ELISpot in a subset of 24 subjects immunized with PHH-1V after a primary vaccination with ChAdOx1-S (Vaxzevria; AstraZeneca). Overall, the heterologous boost with the PHH-1V vaccine, after a primary immunization with ChAdOx1-S, elicits a significant increase in RBD/Spike-specific IFN-γ^+^ T-cell response against different SARS-CoV-2 variants tested compared to baseline. Likewise, CD4^+^ and CD8^+^ T cell responses were also analyzed on Day 14 by ICS in the same subset of subjects. In conclusion, the heterologous boost with the PHH-1V vaccine, after a primary immunization with ChAdOx1-S, elicits T-cell responses with a balanced Th1/Th2 profile with activation of RBD/Spike specific CD4^+^ T-cells. Our study was subject to some limitations. Immunogenicity assessment at 3, 6 and 12 months has only been studied in a subset of 235 subjects. Most of the participants were also primed with mRNA vaccines and only a small sample had an adenovirus vaccine shot. It has not been analyzed neutralizing antibodies against current circulation variants (XBB.1.5, XBB.1.16, EG.5.1, BA.2.86) neither T-cells responses against any Omicron variant. Nevertheless, we reported in a recent publication that PHH-1V induces a long-term immune response up to 6 months to XBB.1.5 SARS-CoV-2 variant and it was non-inferior compared with BNT162b2^22^. Furthermore, neutralizing activity was only analyzed by PBNA and not by a live SARS-CoV-2 neutralization assay, despite this, new validation analysis confirms the validity of the PBNA for SARS-CoV-2^41^.

In conclusion, PHH-1V vaccine (BIMERVAX^®^, HIPRA) was well tolerated and safe, regardless of the primary vaccination schedule received or previous SARS-CoV-2 infection. Heterologous booster with PHH-1V induce a broad and long-lasting humoral immune response against different SARS-CoV-2 variants and could be an interesting strategy for upcoming vaccination campaigns in individuals already immunized with mRNA and/or vector vaccines. PHH-1V vaccine received a marketed authorization of the EMA as a booster in people aged 16 years and above who have been vaccinated with an mRNA COVID-19 vaccine^42^.

## Data Availability

All data relevant to the study are included in the article or uploaded as supplementary information. Further data are available from the authors upon reasonable request and with permission of HIPRA S.A.

## CONTRIBUTORS

Veristat was responsible for managing the data and performing the statistical analyses. Authors contributed to the acquisition, analysis, and/or interpretation of data. All authors had full access to all the data, revised the manuscript critically for important intellectual content, approved the version to be published, and accepted responsibility for publication.

## ACKNOWLEDGEMENTS

Medical writing support was provided by Adrian Lázaro-Frías at Evidenze Health España S.L.U during the preparation of this paper, and funded by HIPRA SCIENTIFIC, S.L.U. We especially acknowledge the following members of Veristat, who contributed to the success of this trial. The following were responsible for study management, biostatistics, medical monitoring, data management, and database programming of the study: Lubia Álvarez, MD, Robin Bliss, PhD, Judith Oribe, Emma Albacar, MPH, Nancy Hsieh, MPH, Marcela Cancino, MSc, Rachel Smith, Montse Barcelo, MD, Mariska van der Heijden, MSc, Amy Booth, Edmund Chiu and Rodney Sleith, MS, Avani Patel, Atalah Haun, MD and Cesar Wong, MD.

We are grateful to Daniel Perez-Zsolt and Jordana Muñoz-Basagoiti for their outstanding contribution to VNA.

The authors would like to thank Silvia Marfil, Raquel Ortiz and Carla Rovirosa for technical assistance.

The authors thank Ruth Peña, Esther Jimenez-Moyano for technical assistance with sample management and ELISpot and Gabriel Felipe Rodriguez-Lozano for ELISpot database generation.

We would like to express our gratitude to Marina Machado, Ana Álvarez-Uría, Sara Rodríguez, Mª Jesús Pérez Granda, Juan Carlos López Bernaldo de Quirós, Mª Teresa Aldamiz, Francisco Tejerina, Rocío Fernández, Martha Kestler, Cristina Díez, Iván Adán, Ana Mur, Patricia Gómez, Félix García and Víctor Fernández for their tireless effort and contribution to this important public health clinical trial.

We would like to thank Glòria Pujol and Eduard Fossas for their assistance in the revision of the manuscript; Fiorella Gallo, Núria Fuentes and Miriam Oria for the ELISA analysis; Clara Panosa, Thais Pentinat and Ester Puigvert for their assistance in the production of the vaccine and Jordi Palmada and Eva Pol for carrying out manufacturing controls. And of course, we would like to especially thank all the HIPRA workers who in one way or another have contributed to making this project a reality.

The authors thank the members of the DSMB for their expertise and recommendations. We are indebted to the HCB-IDIBAPS Biobank, integrated in the Spanish National Biobanks Network, for the biological human samples and data procurement. We want to express our gratitude to all the volunteers for their time and effort. With their contribution they have enabled the generation of medical and scientific knowledge that will enable us to draw closer to the end of this pandemic.

## SUPPLEMENTARY INFORMATION

## Supplementary methods

**Table S1.**
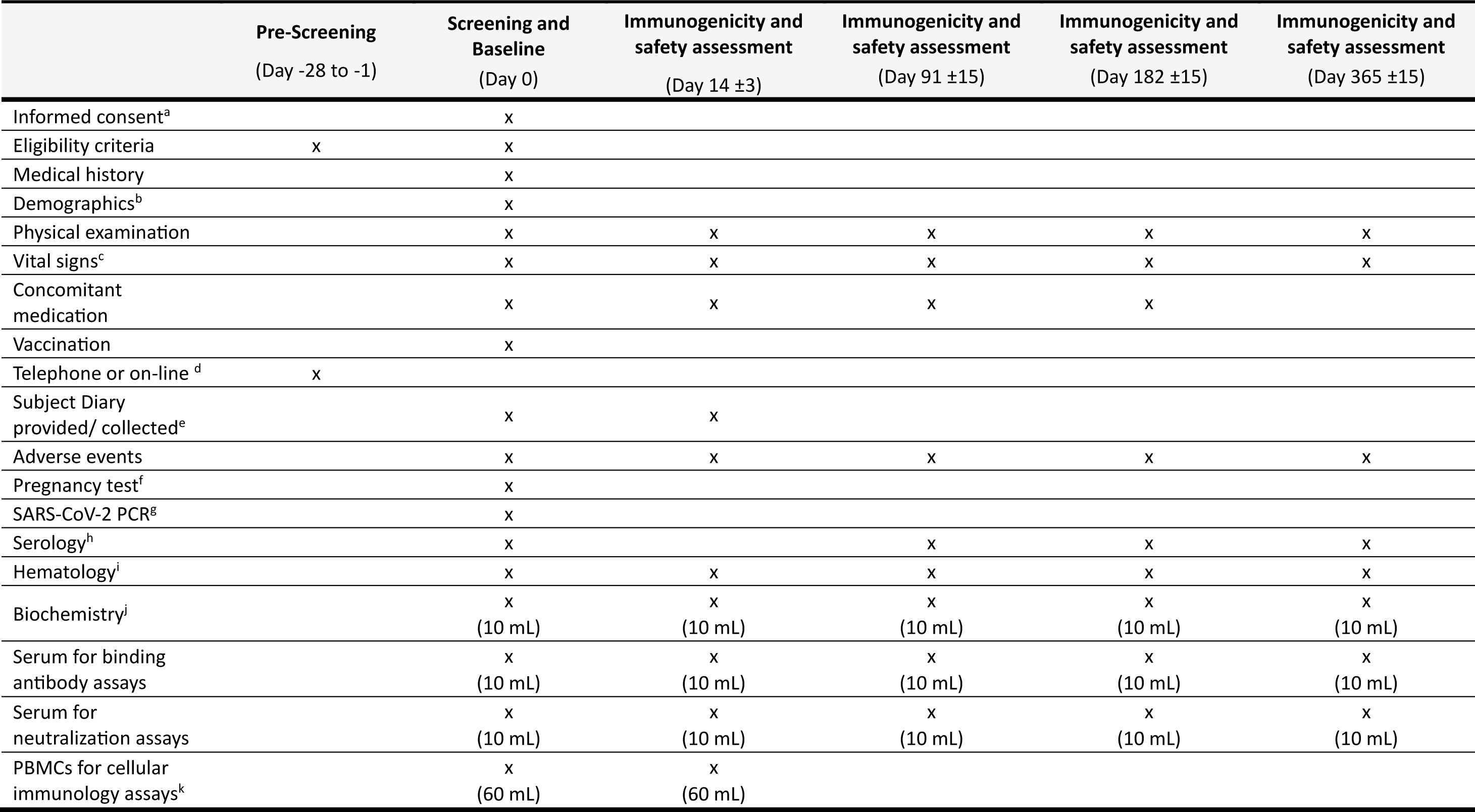
Protocol schedule of the study. ^a^Informed consent was provided via email to the subject after the Pre-Screening and once the study was approved by the ethics committee and competent authorities. ICF was signed and dated at Baseline Vaccine visit. ^b^Demographics included gender, race, ethnicity, and age. ^c^Vital signs included pulse, blood pressure, temperature, and oxygen saturation. ^d^Telephone or on-line visit was performed at Pre-Screening to assess eligibility criteria and to ask for email address to send the ICF. ^e^A paper Subject Diary was provided to all subjects to collect solicited local reactions and systemic events after vaccination. The Subject Diary was collected at the Day 14 visit. ^f^Urine pregnancy test was performed at the study site, only for women of childbearing potential on Day 0. A negative urine pregnancy test should have been verified before vaccination. ^g^SARS-CoV-2 PCR was only performed for subjects with primary vaccination with two doses of Vaxzevria. ^h^Serology was only performed for subjects with primary vaccination with two doses of Vaxzevria to check if they have had a previous COVID-19 infection. ^i^Hematology included CBC, hemoglobin, and platelets. ^j^Biochemistry included alanine aminotransferase, aspartate aminotransferase, gamma-glutamyl transferase, alkaline phosphatase, total bilirubin, BUN, or urea (urea can be measured), creatinine, glucose, potassium, sodium, and total protein. 10 mL of blood was required for hematology and biochemistry. ^k^Only for a subset of 30 subjects with primary vaccination with two doses of Vaxzevria included in the cellular immunogenicity. ***Abbreviations:*** *BUN = blood urea nitrogen; CBC = complete blood count; ETV = early termination visit; ICF = informed consent form; PBMC = peripheral blood mononuclear cells; PCR = polymerase chain reaction; USC = unscheduled visit*.

|  | Pre-Screening<br>(Day -28 to -1) | Screening and<br>Baseline<br>(Day 0) | Immunogenicity and<br>safety assessment<br>(Day 14 ±3) | Immunogenicity and<br>safety assessment<br>(Day 91 ±15) | Immunogenicity and<br>safety assessment<br>(Day 182 ±15) | Immunogenicity and<br>safety assessment<br>(Day 365 ±15) |
| --- | --- | --- | --- | --- | --- | --- |
| Informed consent<sup>a</sup> |  | x |  |  |  |  |
| Eligibility criteria | x | x |  |  |  |  |
| Medical history |  | x |  |  |  |  |
| Demographics<sup>b</sup> |  | x |  |  |  |  |
| Physical examination |  | x | x | x | x | x |
| Vital signs<sup>c</sup> |  | x | x | x | x | x |
| Concomitant<br>medication |  | x | x | x | x |  |
| Vaccination |  | x |  |  |  |  |
| Telephone or on-line <sup>d</sup> | x |  |  |  |  |  |
| Subject Diary<br>provided/ collected<sup>e</sup> |  | x | x |  |  |  |
| Adverse events |  | x | x | x | x | x |
| Pregnancy test<sup>f</sup> |  | x |  |  |  |  |
| SARS-CoV-2 PCR<sup>g</sup> |  | x |  |  |  |  |
| Serology<sup>h</sup> |  | x |  | x | x | x |
| Hematology<sup>i</sup> |  | x | x | x | x | x |
| Biochemistry<sup>j</sup> |  | x<br>(10 mL) | x<br>(10 mL) | x<br>(10 mL) | x<br>(10 mL) | x<br>(10 mL) |
| Serum for binding<br>antibody assays |  | x<br>(10 mL) | x<br>(10 mL) | x<br>(10 mL) | x<br>(10 mL) | x<br>(10 mL) |
| Serum for<br>neutralization assays |  | x<br>(10 mL) | x<br>(10 mL) | x<br>(10 mL) | x<br>(10 mL) | x<br>(10 mL) |
| PBMCs for cellular<br>immunology assays<sup>k</sup> |  | x<br>(60 mL) | x<br>(60 mL) |  |  |  |

**Table S2.**
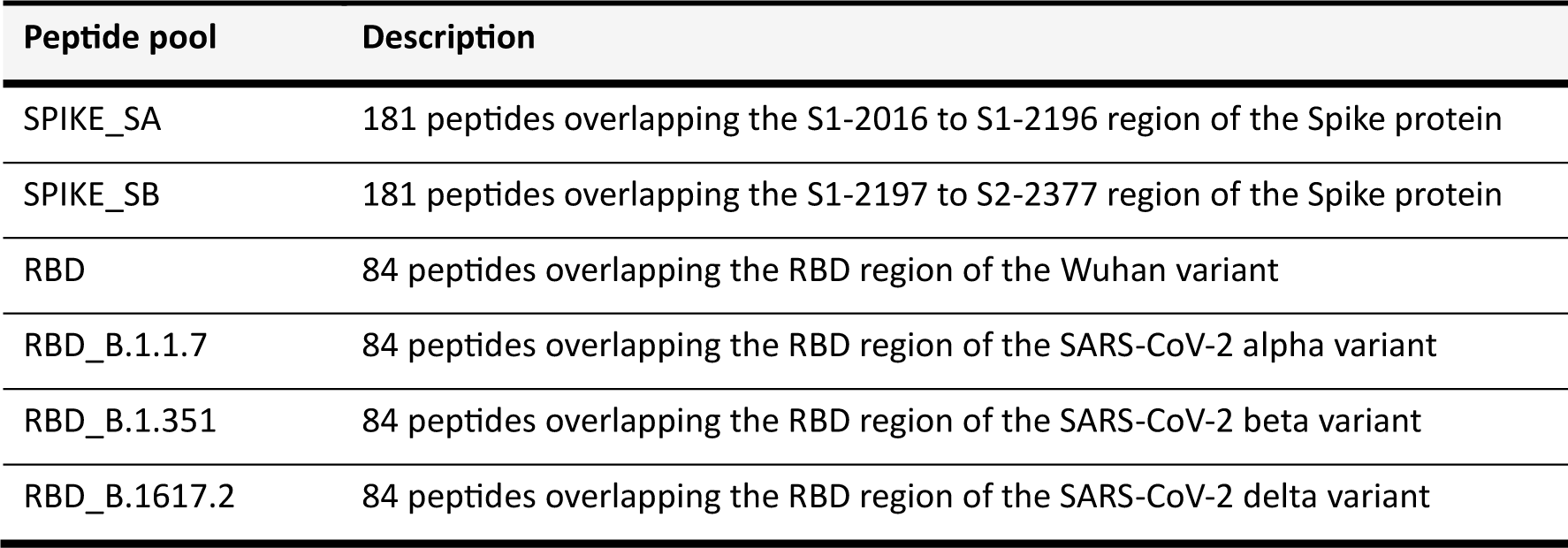
Peptide pools for cellular immunology assays.

| Peptide pool | Description |
| --- | --- |
| SPIKE_SA | 181 peptides overlapping the S1-2016 to S1-2196 region of the Spike protein |
| SPIKE_SB | 181 peptides overlapping the S1-2197 to S2-2377 region of the Spike protein |
| RBD | 84 peptides overlapping the RBD region of the Wuhan variant |
| RBD_B.1.1.7 | 84 peptides overlapping the RBD region of the SARS-CoV-2 alpha variant |
| RBD_B.1.351 | 84 peptides overlapping the RBD region of the SARS-CoV-2 beta variant |
| RBD_B.1617.2 | 84 peptides overlapping the RBD region of the SARS-CoV-2 delta variant |

## Supplementary results

### Safety results

**Table S3.** Summary of Treatment-Emergent Adverse Events (TEAEs) by MedDRA System Organ Class and Preferred Term in ≥1.0% of Subjects. *A TEAE is defined as an adverse event that started on or after the date of administration of study treatment until 28 days thereafter. If a subject experienced more than one TEAE, the subject is counted once for each SOC and once for each PT. SOCs are ordered in decreasing frequency of the total number of subjects with TEAEs reported in each SOC, and PTs are ordered within a SOC in decreasing frequency of the total number of subjects with each TEAE. Adverse events were coded using the MedDRA Dictionary, version 26.0 ***Abbreviations:*** *MedDRA = Medical Dictionary for Regulatory Activities; N = the number of subjects in the population; PT = preferred term; SOC = system organ class; TEAE = treatment-emergent adverse event*.

| System organ class<br>(Preferred term) | PHH-1V (N=2661) |  |
| --- | --- | --- |
|  | Events | Subjects (%) |
| **Total number of TEAEs\*** | **7573** | **2347 (88.20)** |
| **General disorders and administration site conditions** | **5252** | **2276 (85.53)** |
| Injection site pain | 3780 | 2204 (82.83) |
| Fatigue | 851 | 844 (31.72) |
| Injection site swelling | 241 | 241 (9.06) |
| Injection site erythema | 191 | 191 (7.18) |
| Pyrexia | 49 | 49 (1.84) |
| Axillary pain | 38 | 37 (1.39) |
| Injection site induration | 31 | 31 (1.16) |
| **Nervous system disorders** | **892** | **855 (32.13)** |
| Headache | 848 | 831 (31.23) |
| **Musculoskeletal and connective tissue disorders** | **608** | **574 (21.57)** |
| Myalgia | 562 | 552 (20.74) |
| **Gastrointestinal disorders** | **421** | **330 (12.40)** |
| Diarrhea | 207 | 204 (7.67) |
| Vomiting | 157 | 157 (5.90) |
| **Infections and infestations** | **130** | **127 (4.77)** |
| COVID-19 | 53 | 53 (1.99) |
| **Blood and lymphatic system disorders** | **54** | **52 (1.95)** |
| Lymphadenopathy | 33 | 33 (1.24) |

**Table S4.** Summary of Unsolicited Adverse Events from Day 0 through Day 28 in ≥1.0% of Subjects. If a subject experienced more than one event, the subject is counted once for each type of event. ***Abbreviations:*** *COVID-19 = coronavirus disease 2019; N = the number of subjects in the population*.

| Unsolicited AE (≥1.0% of Subjects) | PHH-1V (N=2661) |  |
| --- | --- | --- |
|  | Events | Subjects (%) |
| **Total** | **712** | **538 (20.22)** |
| COVID-19 | 53 | 53 (1.99) |
| Axillary pain | 38 | 37 (1.39) |
| Lymphadenopathy | 33 | 33 (1.24) |
| Injection site induration | 31 | 31 (1.16) |

### Immunogenicity results

**Table S5.** Analysis of neutralizing antibody against SARS-CoV-2 variants after a booster dose with PHH-1 in prime vaccinations groups. Mean GMTs for adjusted treatment and corresponding 95% CI against SARS-CoV-2 Wuhan, Beta, Delta and Omicron BA.1 variants in prime vaccinations groups BNT162b2/BNT162b2 (individuals 16 17 years old; n=13), mRNA-1273/mRNA-1273 (n=172) or ChAdOx1-S/ChAdOx1-S (n=42), or a combination of ChAdOx1-S and another brand of vaccine (n=8, 7 were vaccinated with ChAdOx1-S/BNT162b2and one with mRNA-1273/ChAdOx1-S) at baseline and Days 14, 91, 182 and 364 post-boost are shown. Mean GMFR calculated between time point GMT for adjusted treatment and baseline GMT with GMFR p-value for odds ratio=1 were shown. *p<0,05; **p<0,001; ***p<0,0001. Raw data provided as < 20 have been imputed as 20 for the purposes of analysis. Any zero values have been imputed to 10 (half the LLOQ) for analysis purposes. In all timepoints except Day 365/ETV, raw data provided as > 20480 have been imputed as 20480 for the purposes of analysis. Subjects who reported COVID-19 infections were excluded of this analysis from the report day onwards. ***Abbreviations***: *CI, confidence interval; GMT, geometric mean titer; GMFR, geometric mean fold rise; ETV, early termination visit; LLOQ, lower limit of quantification*.

|  | BNT162b2/BNT162b2<br>(N=13) |  | mRNA-1273/mRNA-1273<br>(N=172) |  | ChAdOx1-S/ChAdOx1<br>(N=42) |  | ChAdOx1-S/ Another<br>(N=8) |  |
| --- | --- | --- | --- | --- | --- | --- | --- | --- |
|  | GMT [95% CI] | GMFR [95% CI] | GMT [95% CI] | GMFR [95% CI] | GMT [95% CI] | GMFR [95% CI] | GMT [95% CI] | GMFR [95% CI] |
| **Wuhan-Hu-1** |  |  |  |  |  |  |  |  |
| Baseline | 682,69 (360.427,1293.106) | - | 660,56 (516.148,845.377) | - | 292,31 (200.5,426.155) | - | 77,34 (34.652,172.632) | - |
| Day 14 | 4034,11 (2129.802,7641.104) | 5,98 (2.577, 13.874)*** | 4420,46 (3454.055,5657.246) | 7,06 (4.881, 10.199)*** | 2080,95 (1427.367,3033.815) | 6,74 (4.043, 11.231)*** | 505,25 (226.363,1127.717) | 7,68 (2.687, 21.940)** |
| Day 91 | 2226,04 (1150.214,4308.12) | 3,27 (1.387, 7.728)* | 1922,8 (1494.222,2474.304) | 3,04 (2.097, 4.416)*** | 714,71 (482.718,1058.196) | 2,22 (1.313, 3.751)* | 405,56 (172.905,951.282) | 6,24 (2.084, 18.696)* |
| Day 182 | 704,35 (343.708,1443.409) | 1,05 (0.424, 2.607) | 1540,9 (1181.807,2009.117) | 2,32 (1.581, 3.390)** | 565,54 (373.051,857.342) | 1,78 (1.033, 3.070) | 861,87 (318.826,2329.864) | 13,82 (4.075, 46.859)*** |
| Day 365 | 937,65 (471.396,1865.092) | 1,39 (0.573, 3.353) | 1117,51 (851.676,1466.312) | 1,67 (1.134, 2.451)* | 335,99 (220.388,512.231) | 1,06 (0.610, 1.828) | 569,81 (190.097,1707.974) | 8,4 (2.266, 31.116)* |
| **Beta** |  |  |  |  |  |  |  |  |
| Baseline | 476,48 (241.313,940.807) | - | 493,64 (398.516,611.47) | - | 488,1 (332.72,716.028) | - | 44,04 (18.635,104.093) | - |
| Day 14 | 7419,1 (3757.433,14649.093) | 13,46 (5.315, 34.112)*** | 6757,78 (5455.561,8370.836) | 13,24 (8.949, 19.599)*** | 4404,52 (3002.431,6461.367) | 8,26 (4.739, 14.408)*** | 1076,66 (455.549,2544.618) | 26,21 (8.201, 83.762)*** |
| Day 91 | 4688,38 (2322.89,9462.733) | 8,58 (3.342, 22.006)*** | 3353,27 (2688.217,4182.862) | 6,61 (4.455, 9.816)*** | 1158,65 (777.051,1727.634) | 2,07 (1.172, 3.647)* | 1588,32 (639.279,3946.255) | 39,54 (11.918, 131.198)*** |
| Day 182 | 3546,72 (1661.679,7570.202) | 6,58 (2.473, 17.530)** | 3817,25 (3012.727,4836.606) | 7,23 (4.836, 10.799)*** | 1738,37 (1138.233,2654.942) | 3,12 (1.745, 5.595)** | 1164,06 (407.086,3328.603) | 26,92 (7.377, 98.276)*** |
| Day 365 | 2054,37 (991.103,4258.311) | 3,63 (1.388, 9.489)* | 1979,35 (1551.577,2525.073) | 3,71 (2.478, 5.566)*** | 748,54 (484.862,1155.597) | 1,32 (0.732, 2.379) | 1042,9 (328.159,3314.338) | 23,49 (6.004, 91.944)*** |
| **Delta** |  |  |  |  |  |  |  |  |
| Baseline | 735,15 (380.029,1422.121) | - | 958,47 (731.498,1255.879) | - | 279,14 (187.958,414.551) | - | 98,13 (42.933,224.289) | - |
| Day 14 | 6103,51 (3155.151,11807.005) | 7,6 (3.080, 18.764)*** | 5991,36 (4572.547,7850.424) | 6,67 (4.552, 9.763)*** | 2398 (1614.693,3561.294) | 8,27 (4.813, 14.199)*** | 949,91 (415.597,2171.159) | 11,42 (3.690, 35.317)*** |
| Day 91 | 3436,09 (1739.327,6788.077) | 4,38 (1.752, 10.971)* | 3904,97 (2965.332,5142.345) | 4,36 (2.966, 6.404)*** | 1292,87 (857.536,1949.213) | 4,24 (2.438, 7.372)*** | 1794,02 (747.028,4308.432) | 21,7 (6.740, 69.865)*** |
| Day 182 | 2525,09 (1207.459,5280.582) | 3,13 (1.202, 8.150) | 1900,65 (1425.79,2533.661) | 2,05 (1.385, 3.033)* | 738,13 (478.407,1138.844) | 2,46 (1.392, 4.348)* | 1032,51 (373.297,2855.833) | 12,02 (3.369, 42.847)** |
| Day 365 | 1369,63 (674.522,2781.042) | 1,64 (0.643, 4.190) | 1203,66 (898.158,1613.079) | 1,29 (0.867, 1.911) | 653,62 (421.298,1014.048) | 2,15 (1.212, 3.813)* | 1149,94 (375.264,3523.818) | 13,29 (3.461, 51.070)** |
| **Omicron BA.1** |  |  |  |  |  |  |  |  |
| Baseline | 288,73 (132.404,629.624) | - | 232,8 (177.76,304.883) | - | 158,45 (101.28,247.885) | - | 37,32 (13.951,99.825) | - |
| Day 14 | 5880,29 (2696.547,12823) | 19,97 (6.995, 57.016)*** | 4508,47 (3442.548,5904.441) | 20,2 (13.508, 30.204)*** | 1638,28 (1047.189,2563.021) | 9,99 (5.421, 18.394)*** | 657,42 (245.769,1758.579) | 18,47 (4.951, 68.874)*** |
| Day 91 | 4429,03 (1982.031,9897.075) | 15,1 (5.209, 43.759)*** | 1676,16 (1270.92,2210.61) | 7,52 (5.009, 11.287)*** | 642,96 (403.615,1024.225) | 3,69 (1.975, 6.888)** | 644,68 (227.801,1824.431) | 18,46 (4.741, 71.910)*** |
| Day 182 | 2319,92 (974.555,5522.545) | 7,49 (2.478, 22.658)* | 1745,03 (1302.087,2338.654) | 7,55 (4.988, 11.431)*** | 858,01 (524.357,1403.957) | 5,03 (2.645, 9.559)*** | 756,97 (228.161,2511.377) | 21,73 (5.000, 94.462)** |
| Day 365 | 1087,91 (472.357,2505.635) | 3,47 (1.171, 10.261) | 959,26 (710.746,1294.666) | 4,11 (2.703, 6.239)*** | 435,52 (264.537,717.021) | 2,53 (1.328, 4.837)* | 509,76 (136.511,1903.559) | 14,95 (3.172, 70.473)* |

**Figure S1.**
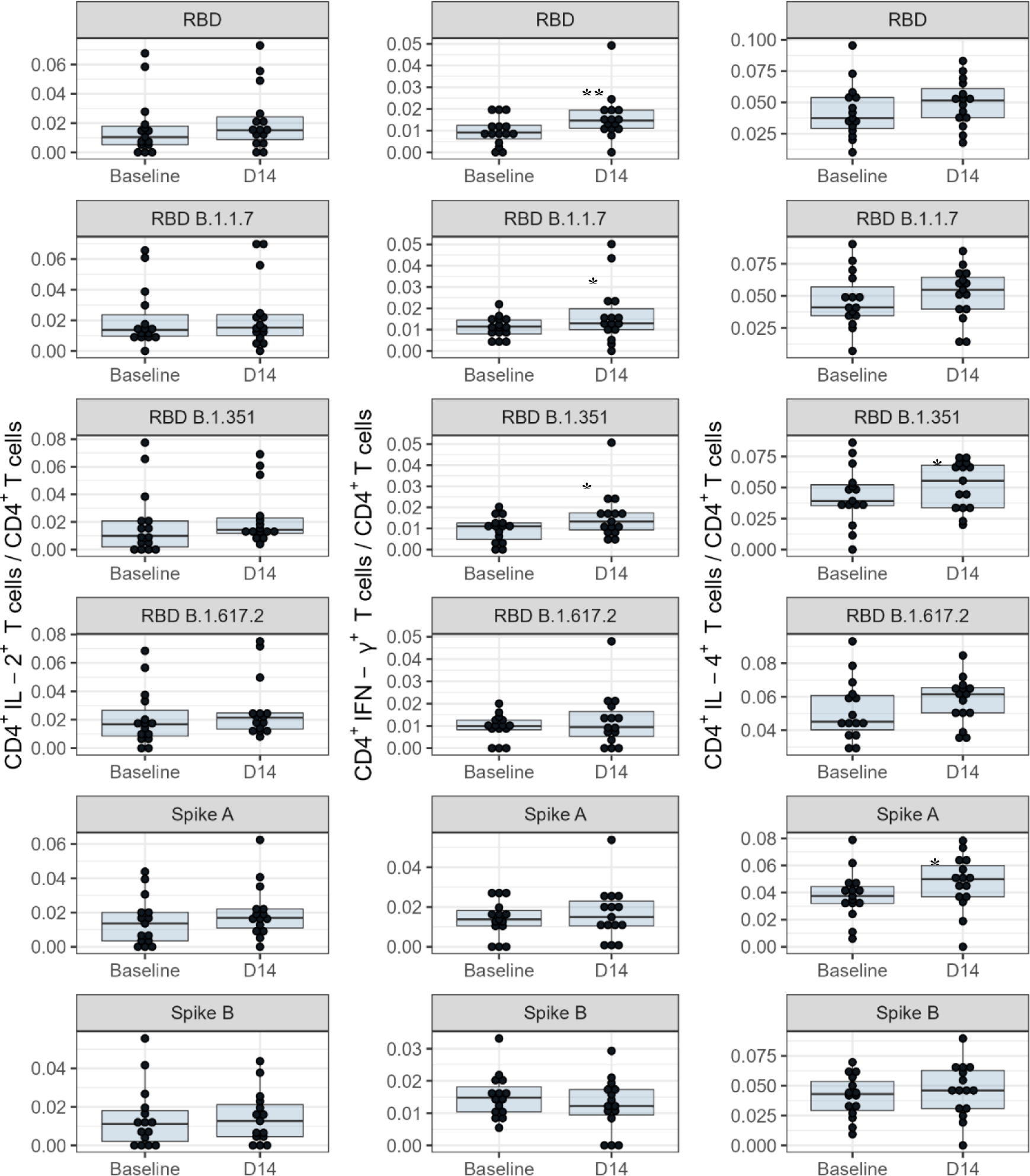
CD4^+^ T-cell responses in PBMC from groups immunized with a PHH-1V booster dose after a primary vaccination with Vaxzevria. PBMC were isolated before the booster vaccination (Baseline) and at Day 14 (Week 2) post vaccination with PHH-1V vaccine, stimulated with RBD (RBD Wuhan, RBD B.1.1.7 or alpha, RBD 1.351 or beta, RBD B.1617.2 or delta) and Spike (SA, SB) peptide pools, and analyzed by intracellular cytokine staining. The frequencies of cytokine expressing CD4^+^ T-cells are shown. The cytokine expression in PBMC stimulated with the medium was considered the background value and this was subtracted from peptide-specific responses. *p<0.05; ** p<0.01. ***Abbreviations:*** *IL-2 = interleukin-2; IL-4 = interleukin-4; IFN-γ = interferon gamma; PBMC = Peripheral Blood Mononuclear Cell; RBD = receptor binding domain*.

**Figure S2.**
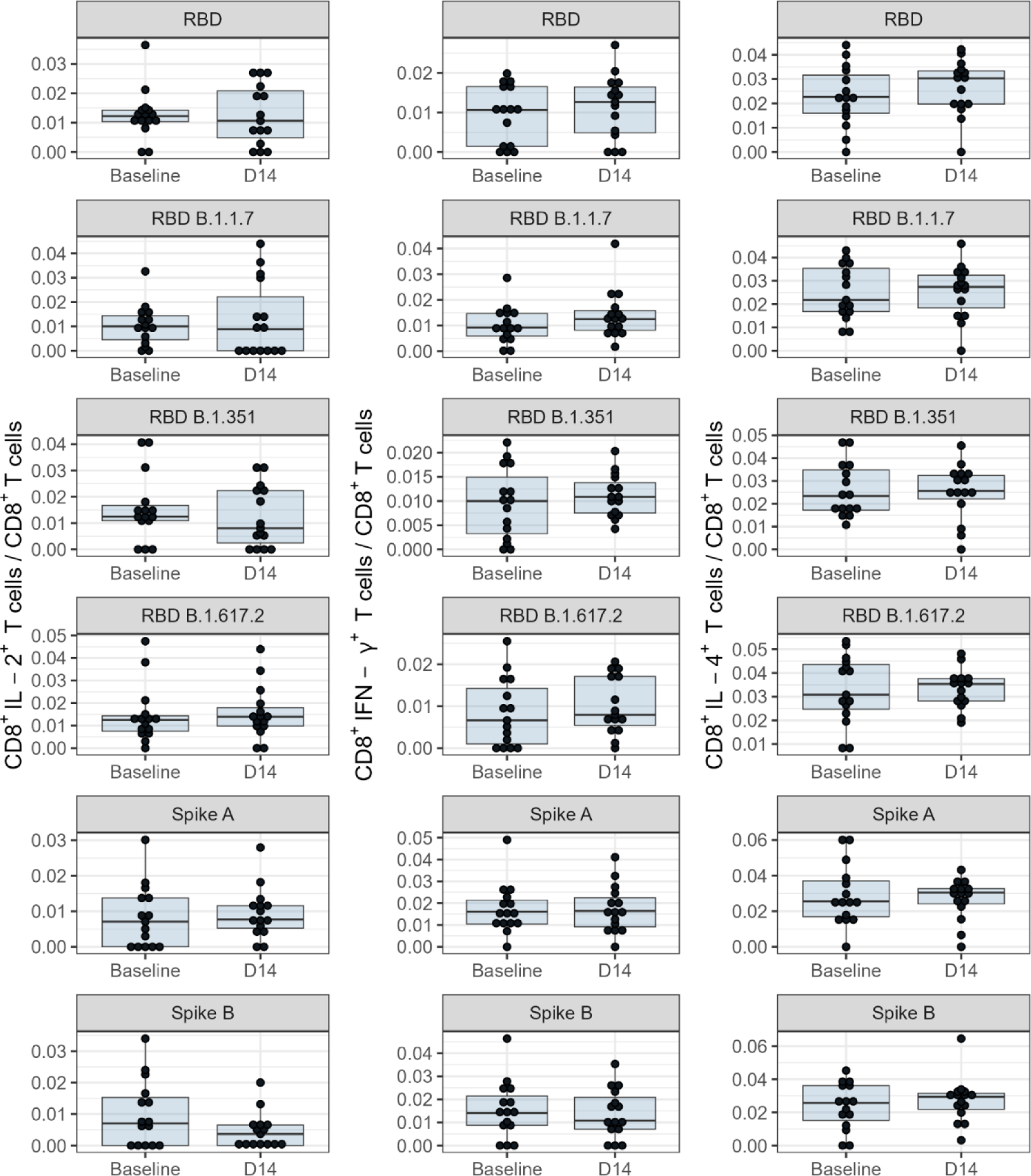
CD8^+^ T-cell responses in PBMC from groups immunized with a PHH-1V booster dose after a primary vaccination with Vaxzevria. PBMC were isolated before the booster vaccination (Baseline) and at Day 14 (Week 2) post vaccination with PHH-1V vaccine, stimulated with RBD (RBD Wuhan, RBD B.1.1.7 or alpha, RBD 1.351 or beta, RBD B.1617.2 or delta) and Spike (SA, SB) peptide pools, and analyzed by intracellular cytokine staining. The frequencies of cytokine expressing CD8^+^ T-cells are shown. The cytokine expression in PBMC stimulated with the medium was considered the background value and this was subtracted from peptide-specific responses. ***Abbreviations:*** *IL-2 = interleukin-2; IL-4 = interleukin-4; IFN-γ = interferon gamma; PBMC = Peripheral Blood Mononuclear Cell; RBD = receptor binding domain*.

